# Monoallelic variation in *DHX9*, the gene encoding the DExH-box helicase DHX9, underlies neurodevelopment disorders and Charcot-Marie-Tooth disease

**DOI:** 10.1101/2023.03.01.23286475

**Authors:** Daniel G. Calame, Tianyu Guo, Chen Wang, Lillian Garrett, Angad Jolly, Moez Dawood, Alina Kurolap, Noa Zunz Henig, Jawid M. Fatih, Isabella Herman, Haowei Du, Tadahiro Mitani, Lore Becker, Birgit Rathkolb, Raffaele Gerlini, Claudia Seisenberger, Susan Marschal, Jill V. Hunter, Amanda Gerard, Alexis Heidlebaugh, Thomas Challman, Rebecca C. Spillmann, Shalini N. Jhangiani, Zeynep Coban-Akdemir, Seema Lalani, Lingxiao Liu, Anya Revah-Politi, Alejandro Iglesias, Edwin Guzman, Evan Baugh, Nathalie Boddaert, Sophie Rondeau, Ormieres Clothide, Giulia Barcia, Queenie K.G. Tan, Isabelle Thiffault, Tomi Pastinen, Kazim Sheikh, Suur Biliciler, Davide Mei, Federico Melani, Vandana Shashi, Yuval Yaron, Mary Steele, Emma Wakeling, Elsebet Østergaard, Lusine Nazaryan-Petersen, Network Undiagnosed Diseases, Francisca Millan, Teresa Santiago-Sim, Julien Thevenon, Ange-Line Bruel, Christel Thauvin-Robinet, Denny Popp, Konrad Platzer, Pawel Gawlinski, Wojciech Wiszniewski, Dana Marafi, Davut Pehlivan, Jennifer E. Posey, Richard A. Gibbs, Valerie Gailus-Durner, Renzo Guerrini, Helmut Fuchs, Martin Hrabě de Angelis, Sabine M. Hölter, Hoi-Hung Cheung, Shen Gu, James R. Lupski

## Abstract

DExD/H-box RNA helicases (DDX/DHX) are encoded by a large paralogous gene family; in a subset of these human helicase genes, pathogenic variation causes neurodevelopmental disorder (NDD) traits and cancer. *DHX9* encodes a BRCA1-interacting nuclear helicase regulating transcription, R-loops, and homologous recombination and exhibits the highest mutational constraint of all DDX/DHX paralogs but remains without disease trait associations. Using exome sequencing and family-based rare variant analysis, we identified 20 individuals with *de novo*, ultra-rare, heterozygous missense or loss-of-function (LoF) *DHX9* variant alleles. Phenotypes ranged from NDDs to the distal symmetric polyneuropathy axonal Charcot-Marie-Tooth disease (CMT2). Quantitative HPO analysis demonstrated genotype-phenotype correlations with LoF variants causing mild NDD phenotypes and nuclear localization signal (NLS) missense variants causing severe NDD. We investigated *DHX9* variant-associated cellular phenotypes in human cell lines. Whereas wild-type DHX9 restricted to the nucleus, NLS missense variants abnormally accumulated in the cytoplasm. Fibroblasts from a patient with an NLS variant also showed abnormal cytoplasmic DHX9 accumulation. CMT2-associated missense variants caused aberrant nucleolar DHX9 accumulation, a phenomenon previously associated with cellular stress. Two NDD-associated variants, p.(Gly411Glu) and p.(Arg761Gln), altered DHX9 ATPase activity. The severe NDD-associated variant p.(Arg141Gln) did not impact DHX9 localization but instead increased R-loop levels and double-stranded DNA breaks. *Dhx9*-/-mice exhibit hypoactivity in novel environments, tremor, and sensorineural hearing loss. Taken together, these results establish *DHX9* as a critical regulator of mammalian neurodevelopment and neuronal homeostasis.

## Introduction

The DExD/H-box (DDX/DHX) gene family consists of 58 highly conserved paralogs encoding RNA helicases^1^. Conserved from bacteria to humans, these proteins share helicase core domains with a consensus DExD or DExH amino acid sequence within the Walker B motif. Despite their evolutionary origin from whole-genome duplication and tandem amplification, DDX/DHX genes are nonredundant and are often essential in model organisms and human cell lines. While broadly implicated in RNA metabolism, the precise function of most DDX/DHX helicases remains unknown, and most lack human disease trait associations^1^. The genes for these helicases are also mutated or dysregulated in cancer and can have oncogenic or tumor suppressive effects^2^.

A growing body of evidence links germline pathogenic variation in DDX/DHX genes to neurodevelopmental disorders (NDD). The X-linked gene *DDX3X* is a leading cause of developmental delay/intellectual disability (DD/ID) in females (MIM: 300958)^3–5^. Other NDD-associated DDX/DHX genes include *DHX30* (MIM: 617805)^6(p30)^, *DDX6* (MIM: 600326)^7(p6)^, and *DDX11* (MIM: 613398)^8^. A large-scale paralog study subsequently provided further evidence for helicase involvement in human brain development as evidenced by identification of four additional DDX/DHX NDD genes, *DHX37* (MIM: 618731), *DHX16* (MIM: 618733), *DDX54*, and *DHX34*, and three candidate NDD genes, *DDX47*, *DHX58*, and *DHX8*^1^.

Among DDX/DHX helicases without disease associations, the DExH-box helicase 9 (DHX9) is particularly intriguing^9^. DHX9 primarily localizes to the nucleus where it regulates transcription and unwinds nucleic acid structures like R-loops, three-stranded structures consisting of a DNA-RNA hybrid and displaced single-stranded DNA^9, 10^. R-loops have a complex role in cell biology, regulating DNA methylation, gene expression and transcription yet also causing single-and double-strand DNA breaks (SSBs and DSBs) with resultant genomic instability^11^. In addition, DHX9 plays an integral role in DNA break repair through homologous recombination (HR) via its recruitment of BRCA1 to DSBs^12^. Dysregulation of transcription, R-loops and SSB/DSB repair are recognized disease mechanisms in neurodevelopmental and neurodegenerative disorders^13, 14(p1),15, 16^. We therefore hypothesized DHX9 dysfunction underlies at least one or more neurologic rare disease traits.

Here, we describe 20 unrelated individuals with sporadic neurologic diseases found to have heterozygous, ultra-rare, *de novo* missense or predicted loss-of-function (pLoF) variant alleles in *DHX9*. Molecular, clinical, and quantitative phenotypic analyses characterized two *DHX9*-associated disease traits, DD/ID and axonal Charcot-Marie-Tooth disease (*i.e*., the distal symmetric polyneuropathy Charcot-Marie-Tooth type 2, CMT2). Cell-based functional studies demonstrate that pathogenic *DHX9* variants cause abnormal cellular distribution of DHX9 and in some cases alter helicase ATPase activity. The impact of DHX9’s loss on organismal biology was studied by generating and phenotyping *Dhx9*^-/-^ mice which exhibit multiple behavioral, neurological, and growth abnormalities. Mechanisms by which *DHX9* variant alleles disrupt neurodevelopment and neuron axonal integrity are explored.

## Subjects and Methods

### Participant Identification and Recruitment

This study was approved by the institutional review board (IRB) at Baylor College of Medicine (BCM) (H-29697). All individuals or their guardians provided written informed consent under BCM protocol H-29697 or through other collaborative IRBs to participate in this study and for the results of this research work to be published. Participants were identified either through the Baylor-Hopkins Center for Mendelian Genomics (BHCMG)/BCM Genomics Research Elucidates the Genetics of Rare disease (BCM-GREGoR) database, the Baylor Genetics (BG) clinical diagnostic laboratory database, GeneMatcher^17, 18^, other research and clinical diagnostic laboratories, or literature search^19, 20^. All subjects were examined by a clinical geneticist and/or neurologist. Pedigrees and deep phenotypic data for each subject were collected from collaborating clinicians using a standardized template. Brain magnetic resonance images (MRI) were collected whenever possible and reviewed by a board-certified neuroradiologist (J.V.H.). Patient were assigned de-identified IDs by the research group which were not made available to anyone outside the research group.

### Exome/Genome Sequencing

Exome sequencing (ES was performed at Baylor College of Medicine Human Genome Sequencing Center (BCM-HGSC) using an Illumina dual indexed, paired-end pre-capture library per manufacturer protocol with modifications as previously described for Subjects 5 (BAB12399), 16 (BAB14692), 17 (BAB704), and BAB4646 and M42-1 (https://www.hgsc.bcm.edu/content/protocols-sequencing-library-construction)21,22. Libraries were pooled and hybridized to the HGSC VCRome 2.1 plus custom Spike-In design according to the manufacturer’s protocol (NimbleGen) with minor revisions^23^. Paired-end sequencing was performed with the Illumina NovaSeq6000 platform. Samples achieved 98% of the targeted exome bases covered to a depth of 20x or greater and had a sequencing yield of 13.2 Gb. Illumina sequence analysis was performed using the HGSC HgV analysis pipeline which moves data through various analysis tools from the initial sequence generation on the instrument to annotated variant calls (SNPs and intra-read in/dels)^24, 25^. In parallel to the exome workflow a SNP Trace panel was generated for a final quality assessment. This included orthogonal confirmation of sample identity and purity using the Error Rate In Sequencing (ERIS) pipeline developed at the BCM-HGSC^26^. Using an “e-GenoTyping” approach, ERIS screens all sequence reads for exact matches to probe sequences defined by the variant and position of interest. A successfully sequenced sample must meet quality control metrics of ERIS SNP array concordance (>90%) and ERIS average contamination rate (<5%).

Subject 4 underwent ES at the the Genetics Institute and Genomics Center, Tel Aviv Sourasky Medical Center using the NovaSeq 6000 platform with IDT xGen Exome Research Panel v2 (Integrated DNA Technologies, Coralville, IA, USA) for library preparation. Reads alighed to GRCh37/hg19. The Franklin by Genoox data analysis platform is used for bioinformatic pipeline and variant analysis.

Subjects 6, 7, and 11 underwent ES at GeneDx (Gaithersburg, MD, USA). Using genomic DNA from the proband and when available, the parents, the exonic regions and flanking splice junctions of the genome were captured using the SureSelect Human All Exon V4 (50 Mb) or the IDT xGen Exome Research Panel v1.0 (Integrated DNA Technologies, Coralville, IA). Massively parallel (NextGen) sequencing was done on an Illumina system with 100bp or greater paired-end reads. Reads were aligned to human genome build GRCh37/UCSC hg19 and analyzed for sequence variants using a custom-developed analysis tool. Reported variants were confirmed, if necessary, by an appropriate orthogonal method in the proband and, if submitted, in selected relatives^27^.

Genome sequencing (GS) was performed for subject 13 and her parents using Nextera DNA Flex library preparation kit, and the libraries were 150 bp paired-end sequenced on a Novaseq 6000 (Illumina, San Diego, CA, USA) following the manufacturers’ instructions. The generated sequencing data had a minimum 10x coverage at least in 98% of mappable positions and an average coverage of 30x. Sequenced reads were trimmed and aligned to the human reference genome GRCh38. Data was processed in accordance with the GATK best practice through GATK v.4.1. For further variant filtering, prioritization and interpretation, VarSeq v.2.2.3 software was used (Golden Helix Inc., Bozeman, MT, United States).

All ES/GS data generated by the BHCMG/BCM-GREGoR for which informed consent for deposition into controlled-access databases was provided was deposited into either dbGap under the BHCMG dbGaP Study Accession phs000711.vs.p1 or into the AnVIL repository under study name Baylor-Hopkins Center for Mendelian Genomics.

### Primary Analysis

The personal genome variation from 12,266 individuals within the BHCMG/BCM-GREGoR database and from 17,500 individuals in the BG clinical diagnostic laboratory database were analyzed for rare, predicted damaging variants in *DHX9*. Variant prioritization utilized minor allele frequencies in gnomAD and the BHCMG/BCM-GREGoR database, conservation (phylop100way, GERP), and functional predictions (MutationTaster, Sorting Intolerant from Tolerant [SIFT], likelihood ratio test [LRT], Combined Annotation Dependent Depletion [CADD], REVEL, SpliceAI, and Human Splice Finder). *De novo* variants were identified from trio ES using DNMFinder^21^. Copy number variant analysis of ES data was performed using XHMM and HMZDelFinder^28, 29^. Candidate variants from ES were orthogonally confirmed by Sanger di-deoxy sequencing.

### Human Phenotype Ontology (HPO) Analysis

Proband phenotypes were annotated with Human Phenotype Ontology (HPO) terms according to their clinically observed findings and then quantitatively analyzed by pairwise similarity between probands calculated using the Lin method^30–32^. This was performed using the OntologyX suite of packages in R^33^. Probands were then clustered according to their phenotypic similarity scores by calculating distance matrices and performing hierarchical agglomerative clustering using the Ward method. The numbers of clusters to consider was defined using a gap statistic curve, and k = 4 was chosen based on the change in slope at that point. Heatmaps were generated using the ComplexHeatmap package in R^34^.

### Expression plasmids

EGFP coding sequence was PCR amplified and cloned into a pCMV5-FLAG-DHX9 vector (Sino Biological) using KpnI to generate the EGFP-tagged DHX9 construct, which expresses the wild type (WT) DHX9 protein. EGFP empty vector was subsequently generated using restriction enzymes KpnI and NotI. Plasmids with DHX9 variants were generated based on EGFP-tagged WT DHX9 vector via recombination using the ClonExpress Ultra One Step Cloning Kit (Vazyme). All plasmid constructs were verified by Sanger sequencing.

### Cell culture and transfection

MCF-7 (human breast cancer cells), PC-3 (human prostate cancer cells) and HEK293T (human embryonic kidney cells) were cultured in the Dulbecco’s Modified Eagle’s Medium (DMEM) supplemented with 10% fetal bovine serum (FBS). Cells were transfected with expression plasmids using Lipofectamine 3000 (Invitrogen) or Lipo 8000 (Beyotime) according to the manufacturers’ instructions. Cells were routinely tested to confirm the absence of mycoplasma contamination. Fibroblasts were generated from patient and paternal skin biopsy and were cultured in DMEM supplemented with 20% FBS.

### Immunocytochemistry

MCF-7, PC-3, and fibroblasts were fixed for 10 min at room temperature with 4% paraformaldeyde, permeabilized with 0.1% Triton X-100 in PBS, and blocked with the blocking buffer (10% Goat Serum, 1% BSA, 0.1% Tween 20). Cells were then incubated overnight with primary antibodies at 4 °C according to the manufacturers’ recommendations; primary antibodies included the anti-Fibrillarin/U3 RNP Rabbit pAb (ABclonal, A13490), anti-DNA-RNA Hybrid Antibody clone S9.6 (Merckmillipore, MABE 1095), anti-RNA helicase A antibody [EPR13521] (Abcam, ab238985), and anti-gamma H2A.X (phospho S139) antibody [EP854(2)Y] (Abcam, ab81299). Post primary antibody incubation, cells were washed in PBS, followed by incubation with mouse Alexa Fluor 647 secondary antibody (Cell Signaling, 4410S) or rabbit Alexa Fluor 555 secondary antibody (Invitrogen, A-21428) for 1 h at 37 °C. Nuclei were stained with 1mg/mL DAPI (Solarbio, C0060). Images were acquired using a confocal microscope (Leica TCS SP8).

### ATPase Assay

HEK293T cells expressing EGFP-tagged DHX9 proteins were purified using the GFP-Trap Agarose Kit (Chromotek) following manufacturer’s instructions. Transfected HEK293T cells were lysed in 200 µL ice-cold radioimmunoprecipitation assay (RIPA) buffer supplemented with DNaseI (75-150 Kunitz U/mL), MgCl^2^ (2.5 mM), protease inhibitor cocktail and phenylmethylsulfonyl fluoride (PMSF) (1 mM), and cell lysates were purified by centrifuging at 17,000x g for 10 min at 4 °C. Proteins were extracted from the supernatant by binding to 20 µL agarose bead slurry for 1 h at 4 °C. Precipitates were first washed twice in washing buffer (10 mM Tris/Cl pH 7.5, 150 mM NaCl, 0.05 % NonidetTM P40 Substitute, 0.5 mM EDTA and 0.018 % sodium azide), and then washed twice in a phosphate-free buffer (40_mM KCl, 35_mM HEPES pH_7.5 and 5_mM MgCl2). Next, precipitates were incubated in a 50 μL phosphate-free reaction mixture containing 2_mM ATP, 2_mM DTT, and 100_μg/ml yeast RNA for 30_min at 30_°C. The amount of free phosphate released by ATP hydrolysis in the ATPase assay was determined by the Biomol Green reagent (Enzo Life Sciences). For each independent ATPase assay, ATPase activity was calculated by subtracting absorbance values of EGFP backbone control from values of EGFP-tagged DHX9 WT or mutant proteins. Subsequently, the amount of EGFP-tagged DHX9 protein was determined by SDS-PAGE Subsequently, the amount of EGFP-tagged DHX9 protein was determined by SDS-PAGE followed by Coomassie blue staining intensity and ATPase activity was normalized to the amount of purified protein.

### *Dhx9* mouse model generation

The *Dhx9*^-/-^ mouse line (*Dhx9^Tm1b (EUCOMM) Hmgu^*) was constructed using the IMPC ‘knockout first’ targeting strategy at Helmholtz Zentrum München, Germany as follows. *Dhx9*^-/-^ mice were generated by allele conversion of C57BL/6NCrl-*Dhx9^Tm1a (EUCOMM) Hmgu^* mouse line originating from EUCOMM ES clone HEPD0554_5_E05 (clone construction overview here: https://www.mousephenotype.org/data/genes/MGI:108177#order). The *tm1b* allele was produced by deletion of exon 4 of *Dhx9* and the neomycin cassette using a cell-permeable Cre recombinase. The allele is a knockout as skipping over of the LacZ cassette does not produce a functional protein. The cassette expresses LacZ under the control of the *Dhx9* promoter as fusion protein with exon 3. The mice were genotyped to verify the mutation and heterozygous mice were intercrossed to generate *Dhx9*^-/-^ mice with *Dhx9*^+/+^ controls for experimental analysis. Mice from five cohorts were used in the analysis to have the following number of mutant mice per group: n = 6 male mutants, n = 9 female mutants. There was no evidence of subviability in the line and the *Dhx9* mice can be ordered, and all related information is available through the IMPC website (https://www.mousephenotype.org/data/genes/MGI:108177#order). RNA quality control analysis using RT-PCR of heterozygous brain tissue revealed the fusion of exon 3 with the synthetic cassette, resulting in a null allele. There was one annotated domain left in the mutated protein. Mice were housed in IVC cages with water and standard mouse chow available *ad libitum* according to the European Union directive 2010/63/EU and German Moues Clinic (GMC) housing conditions (http://www.mouseclinic.de). Moreover, all animal care and use in this study met approval by, and complied with, the rules of the district government of Upper Bavaria (Regierung von Oberbayern), Germany.

### Mouse phenotyping

From the age of 8-16 weeks, the *Dhx9*^-/-^ mice were phenotyped systematically in the GMC as described previously^35^ and in accordance with the standardized phenotyping pipeline of the IMPC (IMPReSS: https://www.mousephenotype.org/impress/index, see **Fig. S1** for overview). The testing details described here are for those assays in which we identified alterations relevant for DHX9 function. Homozygous mutant (“-/-“) and wildtype controls (“+/+”) were compared and the number of animals per group and age of testing for the different assays are shown in **Table S1**. Body weight was measured in the different cohorts.

Data generated by the Open Field test, SHIRPA and grip strength were obtained at 8 and 9 weeks of age. The 20-minute Open Field (OF) test was carried out using the ActiMot system (TSE, Germany) as described previously^36^. The arena was made of transparent and infra-red light-permeable acrylic with a smooth floor (internal measurements: 45.5 x 45.5 x 39.5 cm, 200 lux in middle). For neurological analysis a modified SHIRPA protocol was applied^35, 37, 38^ covering general neurobehavioral aspects that were rated with defined rating scales. During observation in the arena, a trained observer categorized gait (normal, abnormal) and recorded the occurrence of tremor during observation. Grip strength was also measured according to our standard protocol^35, 37, 38^.

Sensorimotor gating and recruitment was measured via assessment of the acoustic startle reflex (ASR) and its prepulse inhibition (PPI) at 10 weeks of age with modification to the previously described protocol^39^ and further details can be found here (https://www.mousephenotype.org/impress/ProcedureInfo?action=list&procID=746&pipeID=14). Briefly, the Med Associates Inc. (St. Albans, USA) startle equipment was used with background noise [no stimulus (NS)] set to 65 dB. Basal startle response (S, startle pulse of 110 dB/40 ms white noise) and percent pre-pulse inhibition, %PPI, to four different pre-pulse (PP) intensities (67, 69, 73, 81 dB [2, 4, 8 and 16 dB above background respectively]), 50-ms interval between S and PP) were determined.

At the age of 11 weeks, the mice were housed individually in metabolic homecages (MHCs) for indirect calorimetry analysis. Forward (distance travelled) and vertical (rearing) locomotor activity as well as food intake and body weight loss were measured (TSE, Germany, detailed protocol: https://www.mousephenotype.org/impress/ProcedureInfo?action=list&procID=855&pipeID=14). The measurement commenced five hours before lights off and finished four hours after lights-on the next morning (21 hours in total).

Altered glucose metabolism was determined using the intraperitoneal glucose tolerance test (ipGTT) at the age of 13 weeks. Glucose was administered intraperitoneally (2 g/kg i.p.) after a 16-h withdrawal of food and glucose levels were measured before and at 15, 30, 60, and 120 minutes after glucose injection. Blood glucose levels were assessed in blood collected from the tail vein with the Accu-Chek Aviva Connect glucose analyser (Roche/Mannheim).

Auditory brainstem response (ABR) was measured at the age of 14 weeks in anesthetized mice as described (https://www.mousephenotype.org/impress/ProcedureInfo?action=list&procID=665&pipeID=7).

At the age of 16 weeks, the final blood samples were collected from the retrobulbar vein plexus under isoflurane anaesthesia in Li-heparin coated tubes (Li1000A, Kabe Labortechnik). The samples were centrifuged at 5000xg for 10 minutes at 8°C and plasma separated within one hour of blood collection. Clinical chemistry parameters were measured immediately using an AU480 analyser (Beckman-Coulter) and adapted reagent kits from Beckman-Coulter according to the manufacturer’s instructions, as described previously^40^. The hematology was analysed with a Sysmex XT-2000iV device using 1:5 diluted samples in the capillary mode as described^41^.

### Statistics

Data was analysed using 2-way ANOVA with *post-hoc* Tukey’s to test genotype x sex interaction effects. For the analysis of rearing activity in the MHC over 21 hours and ABR, a three-way ANOVA was used with genotype, sex and time (for rearing) or frequency (for ABR) as independent variables. Linear regression analysis determined how body weight predicted grip strength. Data was statistically analysed using GraphPad Prism version 8 for Windows (GraphPad Software, La Jolla, California, USA). For all tests, a p value < 0.05 was the level of significance and data are presented as mean ± SD or ± SEM. There was no correction for multiple testing performed.

## Results

### Missense and loss-of-function constraint among DDX/DHX genes

The DDX/DHX superfamily consists of 58 paralogous genes underlying at least 15 rare disease traits. Despite their shared domains and overlapping functions, *DDX* and *DHX* genes vary in their tolerance of missense and pLoF variation (**Fig. 1A,B**)^42^. A negative correlation between missense and pLoF constraint is observed within the superfamily (known disease genes, r = -0.82; all genes, r = -0.76). Strikingly, *DHX9* has the greatest missense and pLoF intolerance of all *DDX*/*DHX* genes (missense Z-score = 5.84, pLI=1, LOEUF=0.1) (**Fig. 1B**). Dominant trait-associated *DDX/DHX* genes tend to have higher pLoF and missense constraint than recessive disease trait-associated genes (**Fig. 1A**). Known haploinsufficient disease genes *DDX3X* and *DHX30* are also highly intolerant of pLoF (LOEUF=0.12). Haploinsufficiency as a disease mechanism is highly enriched in genes in the highest LOEUF decile (≤0.268, dashed line)^42^. Similarly, pHaplo and pTriplo scores for *DHX9*, 0.99 and 1.00, suggest *DHX9* is highly sensitive to both copy number loss and gains^43^. For comparison, *DHX30* has pHaplo and pTriplo scores of 0.97 and 1.00.

**Figure 1:**
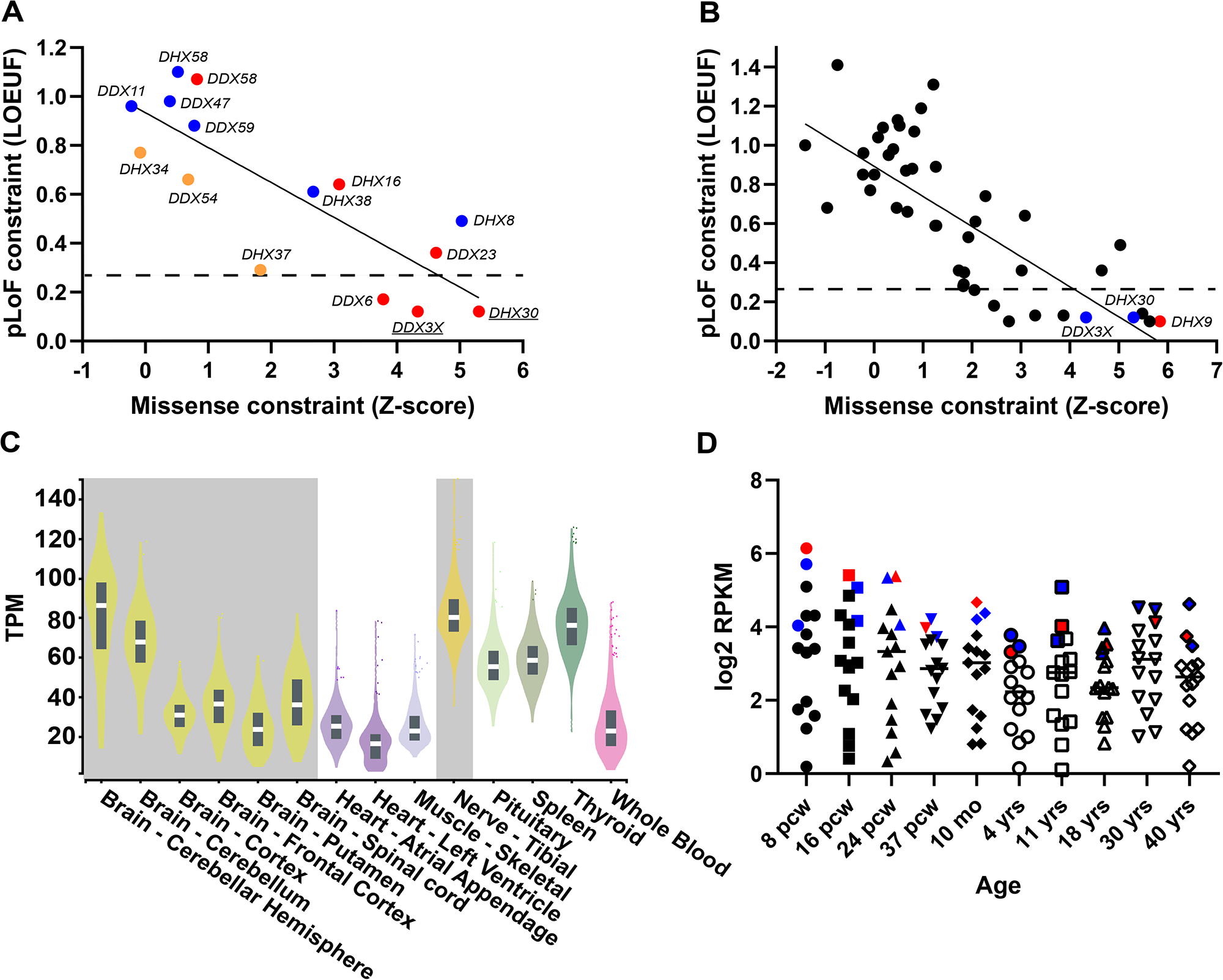
Missense and loss-of-function tolerance in the DDX/DHX superfamily and *DHX9* gene expression. (A) Relationship between pLoF and missense constraint among known or candidate DDX/DHX disease genes. Genes linked to dominant disease traits are shown in red, recessive traits in blue, and mixed dominant/recessive traits in orange. Solid line indicates linear regression. Dashed line shows top LOEUF decile. Y-axis, LOEUF; X-axis, missense Z-score (gnomAD v2.1.1). (B) Relationship between pLoF and missense constraint among all DDX/DHX disease genes. *DHX9* is indicated by a red dot. Paralogs *DHX30* and *DDX3X* are indicated by blue dots. Solid line indicates linear regression. Dashed line shows top LOEUF decile. Y-axis, LOEUF; X-axis, missense Z-score (gnomAD v2.1.1). (C) DHX9 mRNA expression in human adult tissues from the Genotype-Tissue Expression (GTEx) project. Nervous system tissues are highlighted in gray. Y-axis, TPM, transcripts per million. (D) Average mRNA expression of known or candidate DDX/DX disease genes in the developing nervous system from Brainspan. DHX9 is indicated by red dots. Paralogs DHX30 and DDX3X are indicated by blue dots. Y-axis, log2 RPKM, Reads Per Kilobase Million; X-axis, development stage, pcw = post-conception weeks, mo = months, yrs = years.

### Tissue expression and protein-protein interactions of DHX9

Genes underlying a neurologic disease trait should be well-expressed within the adult and/or developing nervous system. We investigated *DHX9* expression levels through human development using GTEx and the BrainSpan Atlas of the Developing Human Brain. DHX9 is robustly expressed in all tissues during adulthood with highest neuronal expression within the cerebellum and tibial nerve. We hypothesized that the large sized Purkinje cells of the cerebellum and long track peripheral nerves, such as the tibial nerve, might be most perturbed for biological homeostasis in postmitotic neurons (**Fig. 1C**). Similarly, DHX9 is among the most abundantly expressed *DDX*/*DHX* disease genes in the developing brain with expression levels comparable to DDX3X and DHX30 (**Fig. 1D**), and single cell expression data from the developing human primary cortex in the UCSC Cell Browser (https://cells.ucsc.edu/) shows DHX9 transcripts are observed in all cell types.

Protein-protein interactions between a gene and known disease genes with shared phenotypes provides supporting evidence for gene disease functional biology. Therefore, we examined the *DHX9* interactome using the STRING database **(Fig. S2**). *DHX9* directly interacts with multiple known neurologic disease trait genes including *EWSR1* (amyotrophic lateral sclerosis, ALS), *AGO2* (Lessel-Kreienkamp syndrome [MIM: 619149]), and *HNRNPU* (developmental and epileptic encephalopathy 54 [MIM: 617391]). Literature review captured other relevant protein-protein interactions involving genes/variant alleles underlying ‘axonopathies’ including *SMN1* (spinal muscular atrophy-1 [MIM: 253300]), *FUS* (ALS 6, with or without frontotemporal dementia [MIM: 608030]), *TAF15* (ALS), and *MATR3* (ALS 21 [MIM: 606070]).

### Identification of patients with candidate disease-causing DHX9 variants

*DDX3X* and *DHX30*-associated NDDs result from *de novo* missense or pLoF variant alleles^3–6, 44^. The observations that *DHX9* shares high missense and pLoF intolerance with *DDX3X and DHX30*, and that all three genes exhibit similar expression profiles in the nervous system, implicate a *DHX9*-associated neurologic disease trait potentially resultant from *de novo*, ultra-rare missense or pLoF variants. Therefore, we analyzed ultra-rare, predicted damaging heterozygous missense or pLoF variants in the 29,766 individuals with ES/GS data within the BHCMG/GREGoR and BG databases and subsequently identified additional cases with *DHX9* variants through the online matchmaker GeneMatcher^17, 18^, DECIPHER^45^, and published NDD cohorts^19, 20^. These efforts uncovered twenty unrelated individuals with candidate disease-causing *DHX9* variants and either NDDs or axonal CMT (**Fig. 2**, **Tables 1 & 2**). We also investigated for individuals with biallelic *DHX9* variants consistent with an autosomal recessive (AR) neurological disease trait but failed to find any compelling biallelic candidate variants.

**Figure 2:**
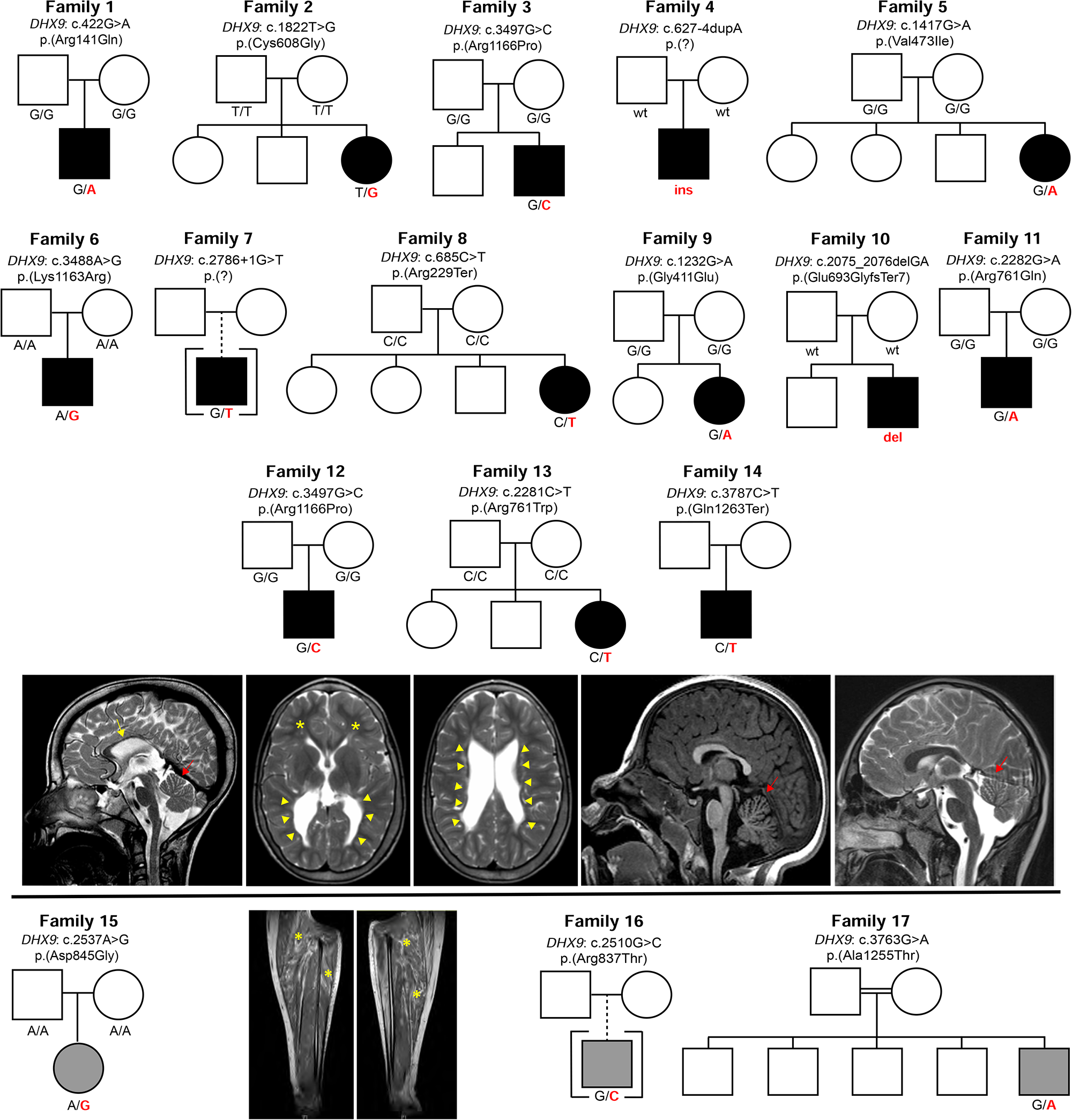
Pedigrees, photographs, and brain imaging of patients with candidate disease-causing *DHX9 variants*. Pedigrees, *DHX9* genotypes, and representative brain MRIs of patients with neurodevelopmental disorders (NDD) are shown above the black line. Patients, genotypes, and representative clinical images and leg muscle MRIs of patients with Charcot-Marie-Tooth disease are shown below the black line. Images of the proband’s feet from Family 15 are redacted per Medrxiv rules regarding identifiable characteristics/body parts but are available upon request from the corresponding author. Yellow arrow indicates thin corpus callosum. Red arrows indicate cerebellar atrophy. Yellow arrowheads show enlargement of the ventricles, and yellow asterisks (brain MRI, second image from right) show reduced white matter volume. Photograph shows pes cavus and hammer toe deformity in Family 15 with CMT. Yellow asterisks (leg MRI) highlight fatty infiltration of the lower leg musculature consistent with CMT. Black pedigree symbols indicate NDDs, whereas gray pedigree symbols indicate CMT.

### Molecular findings

*DHX9* (NM_001357.5) is located on chromosome 1q25.3, contains 28 exons, and encodes a 1270 amino acid (aa) protein (**Fig. 3**). Eight domains of DHX9 have been delineated: two double stranded RNA binding domains (dsRBD), a minimal transactivation domain (MTAD) involved in RNA polymerase II interaction, two helicase domains, a helicase-associated domain 2 (HA2) required for unwinding activity, an oligonucleotide/oligosaccharide-binding fold (OB-fold), and a glycine-rich RGG-box which binds single-stranded nucleic acids^9^. Additionally, there are two nuclear localization signals (NLS): a 9 aa NLS between the helicase domains and a 19 aa NLS within the RGG-box domain. The C-terminal NLS is required for nuclear import via the importin-α3/importin-β pathway^46(p3),47^.

**Figure 3:**
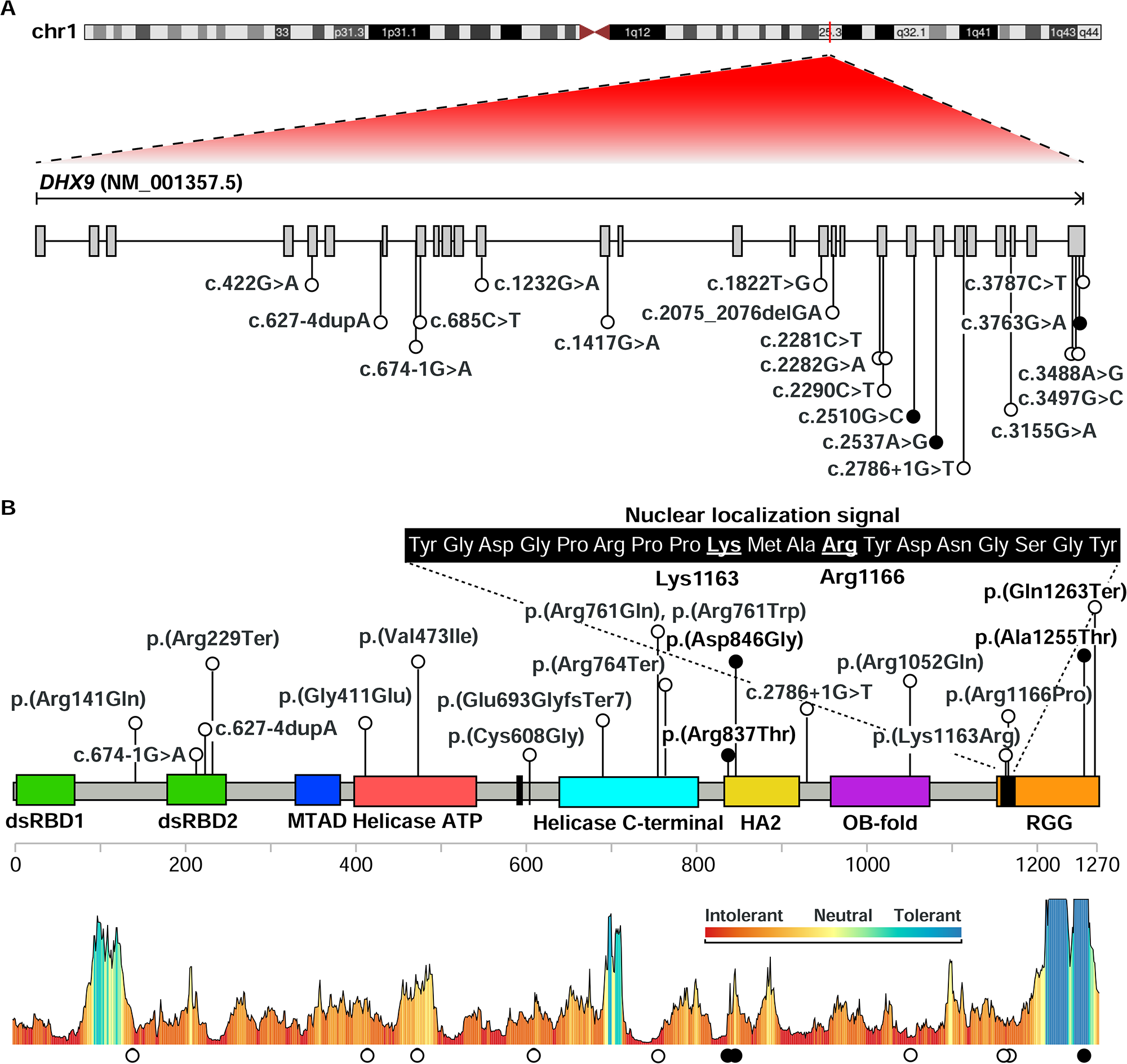
Location of *DHX9 variants*. (A) Diagram of DHX9 mRNA showing location of NDD-associated (white) and CMT-associated (black) variants. (B) Diagram of DHX9 protein showing functional domains including double-stranded RNA-binding domains (dsRBD1&2), minimal transactivation domain (MTAD), helicase domains, helicase associated domain 2 (HA2), oligonucleotide/oligosaccharide-binding fold (OB-fold), and the RGG box. Protein domains were obtained from Uniprot. The sequence of the nuclear localization signal is magnified and the two key residues Lys1163 and Arg1166 are underlined. DHX9’s protein tolerance landscape is shown below the figure as calculated by Metadome.

Variant details, including *de novo* status, CADD and REVEL scores, conservation, and allele frequency are shown as **Table 1**. Nineteen different *DHX9* variants were identified in total including twelve missense variants and five pLoF. All are absent from gnomAD except for c.1417G>A, p.(Val473Ile) (1 heterozygote, allele frequency 4.01e-6). *DHX9* variants occurred *de novo* in fourteen cases: 13 NDD and 1 CMT; in the remaining five cases, parental samples were not available for genotyping. A recurrent *de novo* variant, c.3488A>G, p.(Lys1163Arg), was seen in two patients with sporadic NDDs. Ten missense variants map within functional domains: four within helicase domains, two within HA2 domain, one in OB-fold, and three within RGG-box (**Fig. 3B**). Site-directed mutagenesis previously demonstrated Lys1163 and Arg1166, aa residues impacted by the NLS variants c.3497G>C and c.3488A>G, are required for DHX9 nuclear import^46(p3),47^. Consistent with this, cNLS Mapper, a program which predicts importin -α-dependent NLS^48^, detects the known NLS within DHX9’s C-terminal reference sequence but does not recognize an NLS in either variant sequence (**Fig. S3**). Two of the three CMT-associated *DHX9* variants cluster within the HA2 domain. All *DHX9* missense variants lie in regions with low missense variation tolerance as determined via MetaDome^49^ with the exception of the CMT-associated variant c.3763G>A; p.(Ala1255Thr) (**Fig. 3B**).

**Table 1.** Summary of DHX9 Variant Alleles.

| Individual | Phenotype | Position (hg19) | Nucleotide / Protein | *De novo* | Allele count / frequency (gnomAD) | CADD score | REVEL score | Conservation (phyloP100way) |
| --- | --- | --- | --- | --- | --- | --- | --- | --- |
| 1 | NDD | 1-182822498-G-A | c.422G>A<br>p.(Arg141Gln) | Yes | Absent | 22.3 | 0.1959 | 6.666 |
| 2<br>(BAB15412) | NDD | 1-182845191-T-G | c.1822T>G<br>p.(Cys608Gly) | Yes | Absent | 24.3 | 0.3129 | 7.237 |
| 3, 12 | NDD | 1-182856253-G-C | c.3497G>C<br>p.(Arg1166Pro) | Yes | Absent | 23.8 | 0.6679 | 6.7389 |
| 4 | NDD | 1-182825663-A-AA | c.627-4dupA<br>p.(?) | Yes | Absent | - | - | - |
| 5 | NDD | 1-182835663-G-A | c.1417G>A<br>p.(Val473Ile) | Yes | 1 htz<br>1 in 141,456 | 26.3 | 0.34 | 9.136 |
| 6 | NDD | 1-182856244-A-G | c.3488A>G<br>p.(Lys1163Arg) | Yes | Absent | 22.0 | 0.2669 | 6.17 |
| 7 | NDD | 1-182850561-G-T | c.2786+1G>T<br>p.(?) | Unknown | Absent | 35.0 | - | 9.050 |
| 8 | NDD | 1-182827250-C-T | c.685C>T<br>p.(Arg229Ter) | Yes | Absent | 38 | - | 4.292 |
| 9 | NDD | 1-182829219-G-A | c.1232G>A<br>p.(Gly411Glu) | Yes | Absent | 25.1 | 0.7269 | 9.094 |
| 10 | NDD | 1-182845627-GA- | c.2075_2076delGA<br>p.(Glu693GlyfsTer7) | Yes | Absent | - | - | - |
| 11 | NDD | 1-182847239-G-A | c.2282G>A<br>p.(Arg761Gln) | Yes | Absent | 32 | 0.9039 | 7.328 |
| 13 | NDD | 1-182847238-C-T | c.2281C>T<br>p.(Arg761Trp) | Yes | Absent | 29 | 0.911 | 2.364 |
| 14 | NDD | 1-182856543-C-T | c.3787C>T<br>p.(Gln1263Ter) | Unknown | Absent | 37 | - | 0.829 |
| lossifov *et al.*,<br>2012<sup>1</sup> | NDD | 1-182852665-G-A | c.3155G>A<br>p.(Arg1052Gln) | Yes | Absent | 27.2 | 0.207 | 9.0979 |
| BAB4646<sup>1</sup> | NDD | 1-182827238-G-A | c.674-1G>A<br>p.(?) | Unknown | Absent | 35 | - | 8.947 |
| M42-1<sup>1</sup> | NDD | 1-182847247-C-T | c.2290C>T<br>p.(Arg764Ter) | Unknown | Absent | 36 | - | 1.753 |
| 15<br>(BAB12399) | CMT | 1-182849656-A-G | c.2537A>G<br>p.(Asp846Gly) | Yes | Absent | 26.7 | 0.2099 | 6.8429 |
| 16<br>(BAB14692) | CMT | 1-182848543-G-C | c.2510G>C<br>p.(Arg837Thr) | Unknown | Absent | 31 | 0.1529 | 5.3889 |
| 17<br>(BAB704) | CMT | 1-182856519-G-A | c.3763G>A<br>p.(Ala1255Thr) | Unknown | Absent | 16.19 | 0.1389 | 2.183 |

Abbreviations: NDD, neurodevelopmental disorder; CMT, Charcot-Marie-Tooth disease; CADD, Combined Annotation Dependent Depletion; gnomAD, Genome Aggregation Database; htz, heterozygote; REVEL, rare exome variant ensemble learner; <sup>1</sup> Limited clinical details available or evidence of multilocus pathogenic variation contributing to blended phenotype. See Supplemental Text for additional details.

Five pLoF variants were identified in patients with NDDs: c.2786+1G>T, c.2290C>T; p.(Arg764*), c.685C>T; p.(Arg229*), c.674-1G>A, and c.2075_2076delGA; p.(Glu693GlyfsTer7). All are absent from gnomAD. c.685C>T, c.2290C>T, and c.2075_2076delGA create premature termination codons within exons 8, 18, and 20 of this 28-exon gene, respectively, and therefore are predicted to undergo nonsense-mediated decay (NMD)^50^. c.674-1G>A and c.2786+1G>T lie within introns 7 and 23, respectively, and are predicted to alter splicing by SpliceAI. Other *DHX9* candidate variants include the *de novo* splice variant c.627-4dupA and c.3787C>T; p.(Gln1263Ter). Both are absent from gnomAD. Splicing predictors are divided for c.627-4dupA: Human Splice Finder predicts possible activation of a cryptic acceptor site, whereas SpliceAI does not (**Fig. S4**). c.3787C>T; p.(Gln1263Ter) has a CADD score of 37 and is expected to escape NMD as it lies within the final exon. It should result in a truncated protein lacking the final eight amino acids within the RGG domain.

Since *DHX9* is highly intolerant to pLoF, we also searched our ES database for copy number variants (CNVs) encompassing *DHX9* using XHMM as well as the BG clinical chromosomal microarray database (CMA) and DECIPHER. No small deletion CNVs spanning *DHX9* were detected in the BHCMG/GREGoR database. Similarly, the smallest reported deletion encompassing *DHX9* in the BG CMA database containing ∼90,000 personal genomes was >20 Mb. A 191.52 kb deletion (GRCh37, chr1:182790120-182981637) encompassing *DHX9* and two other coding genes (*NPL*, pLI=0; *SHCBP1L*, pLI=0) was reported in DECIPHER in a subject (ID 288646) with hepatic fibrosis, abnormality of the kidney, global developmental delay, and oculomotor apraxia^45^. The deletion was maternally inherited and classified as ‘likely benign.’ No maternal phenotypic data was provided for review.

### Defining the DHX9-associated disease trait

Clinical records for robust organismal phenotyping of seventeen individuals with candidate disease-causing *DHX9* variants were available. These included data from 14 patients with NDDs and 3 patients with CMT2 used for deep clinical and phenotypic analyses (**Fig. 2**, **Table 2, Supplemental Text**). All individuals with *DHX9*-related NDD had developmental delay (DD) and/or intellectual disability (ID). The degree of cognitive impairment ranged from autism spectrum disorder with speech delay and learning disabilities but normal intelligence (Patient 4) to severe DD/ID (Patients 1, 3, 5, 12). Other developmental or neuropsychiatric disorders were common including anxiety, obsessive-compulsive disorder, autistic spectrum disorders, and neurobehavioral issues (8/14). Other common features included axial hypotonia (7/14) and dysmorphic facial features (8/14) Six individuals had either congenital or postnatal microcephaly (Z-scores -2.14 to -3.39). Seizures were reported in six individuals, and drug-resistant epilepsy occurred in three individuals. Appendicular hypertonia was reported in two individuals. Brain magnetic resonance imaging (MRI) was abnormal in five patients studied. Imaging findings included white matter volume loss with enlargement of the ventricles, corpus callosum thinning, and cerebral and cerebellar atrophy (**Fig. 2**). Other features seen in two or more individuals were cardiac abnormalities (4/14), hyperreflexia (3/14), failure to thrive (2/14), short stature (3/14) and history of recurrent infections (2/14).

**Table 2.**
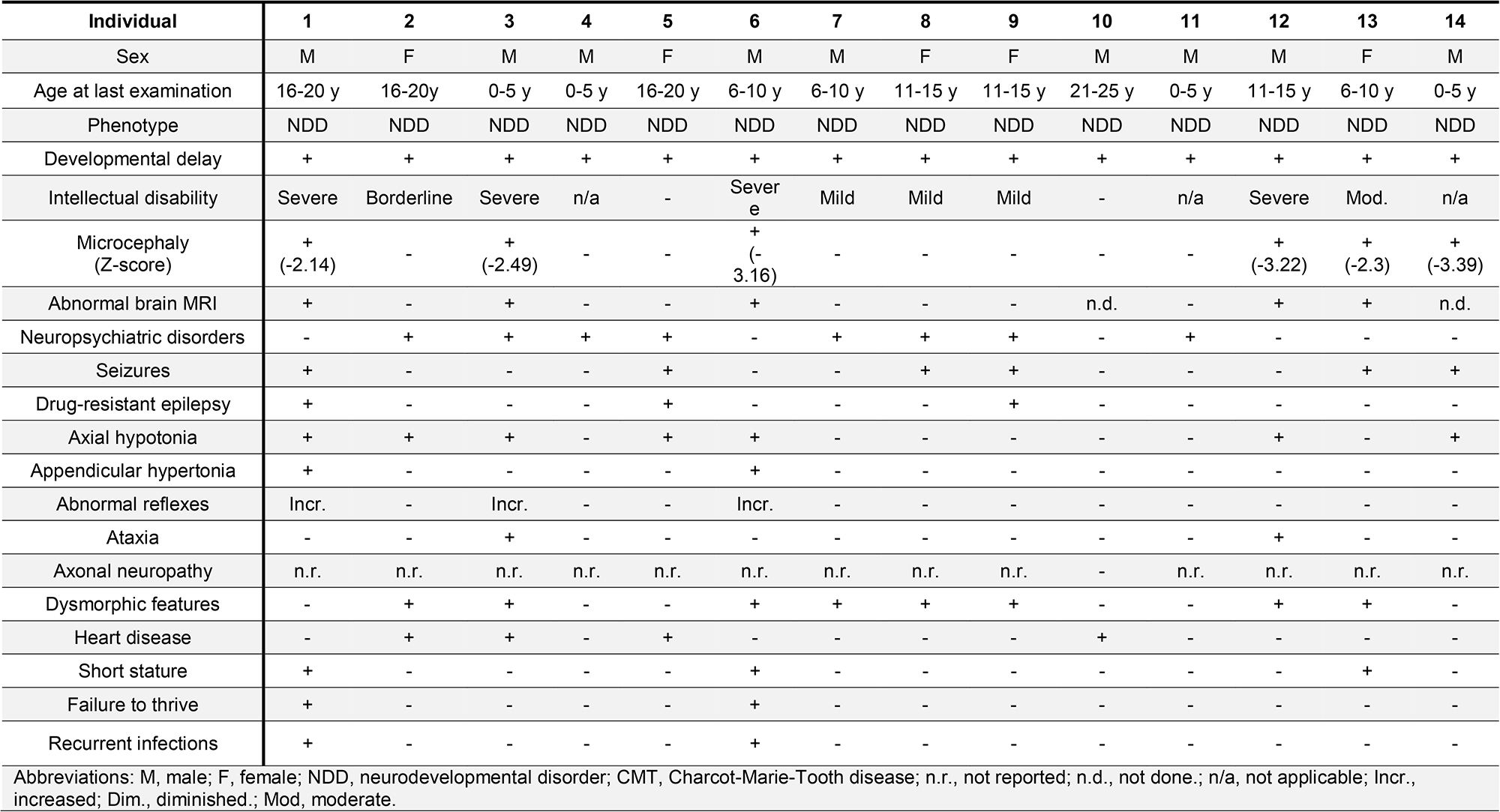
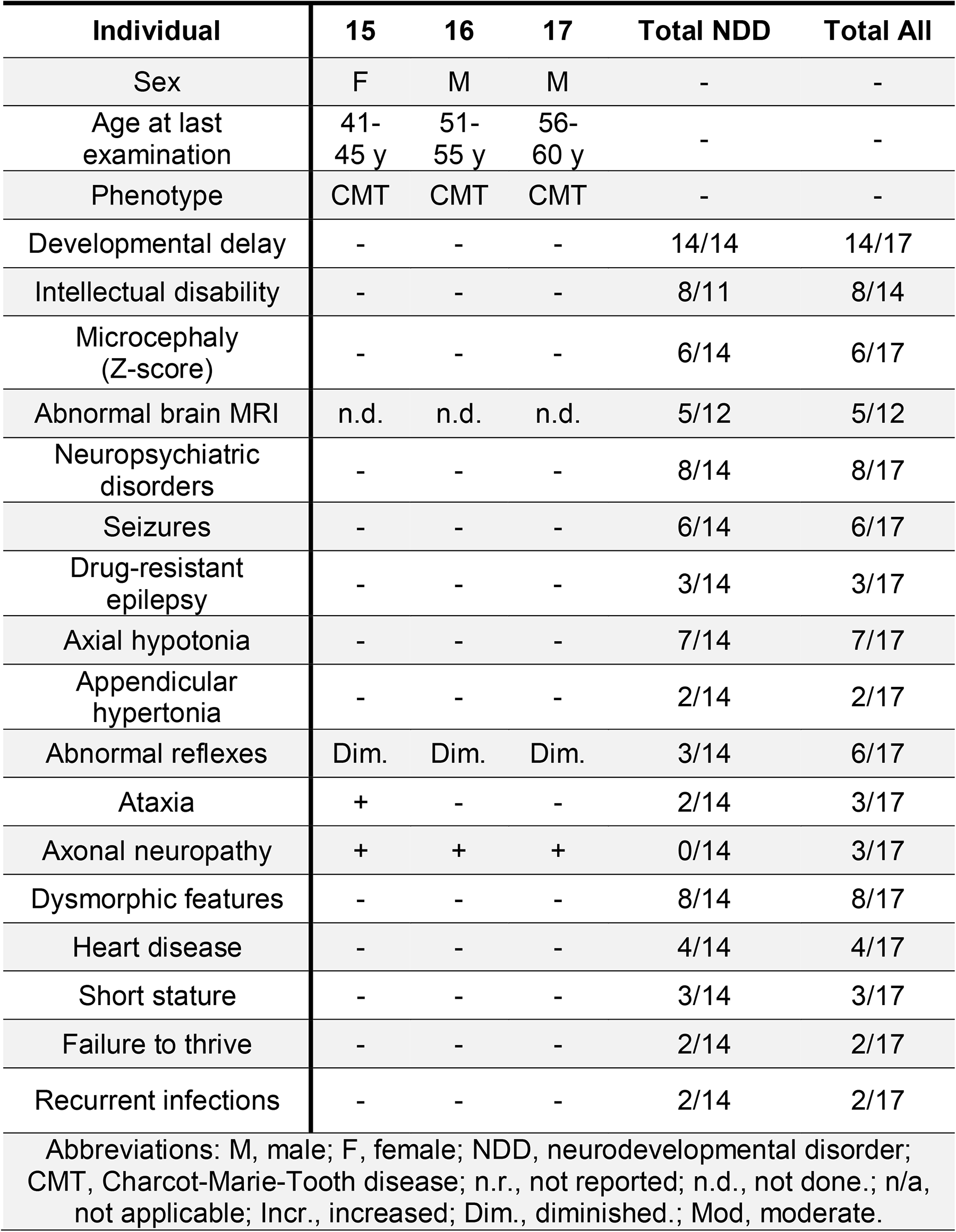
Phenotypic Summary of Individuals with DHX9-related Neurodevelopmental Disorders and CMT.

Three individuals were identified with candidate disease-causing *DHX9* variants and CMT2, also known as hereditary motor and sensory neuropathy (HMSN). Disease onset occurred in adulthood (2/3) or adolescence (1/3). All patients had typical features of CMT, a distal symmetric polyneuropathy (DSP), including distal weakness, sensory deficits, and/or variable muscle wasting or foot deformities (**Fig. 2**)^51^. Two had painful sensory neuropathy.

Electrophysiological features of long tract nerve function were investigated by nerve conduction velocity studies (NCVs). NCVs showed either diminished compound muscle action potential (CMAP) and sensory nerve action potential (SNAP) amplitudes (2/3) or normal nerve conduction velocities (NCVs) and amplitudes (1/3). Electromyography in the patient with normal NCVs showed neurogenic changes in the distal lower extremities and upper extremities and active denervation in the lower extremities. Thus, electrodiagnostic studies supported an axonal neuropathic process in all CMT2 cases.

### Quantitative dissection of genotype-phenotype relationships at the DHX9 locus

Human Phenotype Ontology (HPO), a structured ontology of medical terms and a standardized terminology, is computationally accessible for informatic similarity comparisons of human phenotypic data and rare disease traits; the latter by searchable queries of OMIM clinical synopses. HPO approaches can quantitatively dissect complex disease phenotypes resulting from multilocus pathogenic variation (MPV) and reveal previously unrecognized genotype-phenotype correlations within disease cohorts^52, 53^. The *DHX9* variants encompassing multiple protein functional domains, variant types, and phenotypes were observed to spread over a neurologic disease spectrum; therefore, we performed a quantitative phenotypic similarity analysis of the first fourteen individuals identified without evidence of MPV and for whom detailed phenotypic data were available (**Fig. 4, Supplemental Text**). Phenotypic similarity scores for each proband were calculated and visualized in a cluster heatmap where cluster number was determined by the gap statistic curve (**Fig. 4A**).

**Figure 4:**
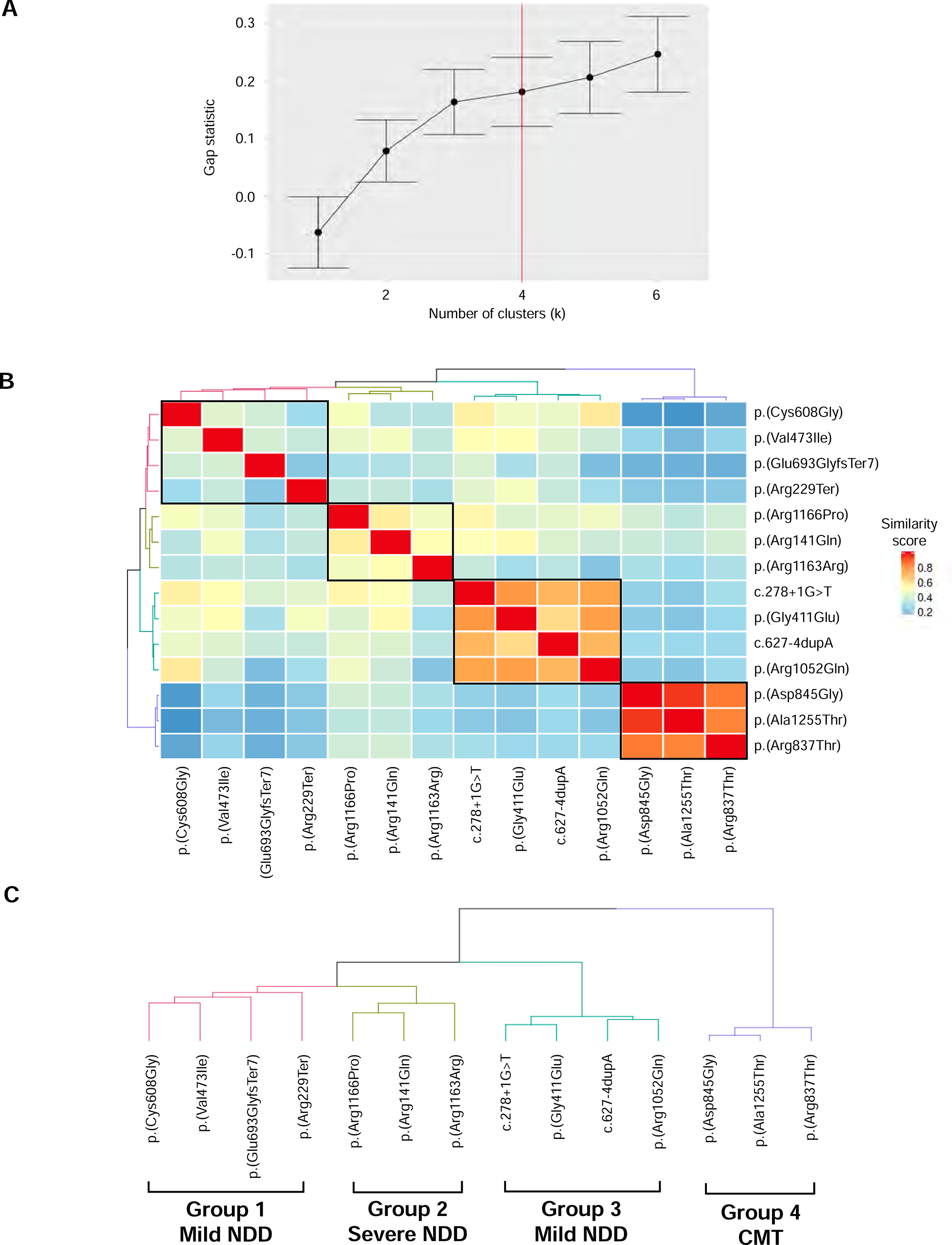
HPO analysis of *DHX9* cohort. (A) Gap statistic curve for *DHX9* cohort. Gap statistic is displayed on the y axis and number of clusters tested on the x axis. The point on the curve where slope changed from a trend of higher to lower (i.e., additional clusters not adding as much to the gap statistic) was chosen as optimal number of clusters (k=4). (B) HAC and visualization of quantitative phenotypic similarity allow refinement of genotype-phenotype correlations in *DHX9* cohort. The dendrogram shown at the top and to the left of the heatmap is based on HAC analysis of the dissimilarity matrix produced from Resnik semantic similarity scores and with k=4. Unique clusters are represented by different colors, and individual probands are labeled on top of and to the right of the heatmap. Within the heatmap, dark red indicates a higher similarity, while dark blue indicates lower similarity. A key is provided on the right. (C) Magnified dendrogram showing unique clusters and group characteristics.

HPO analysis identified four distinct phenotypic clusters (**Fig. 4B, C**). Groups 1 and 3 consisted of individuals with mild NDD phenotypes (*i.e.*, mild deviations from normotypical behaviors such as mild DD/ID, autism spectrum disorders, speech delay) without microcephaly or brain abnormalities, whereas Group 2 contained the most severe phenotypes (severe DD/ID, microcephaly, and brain abnormalities). Group 4 consisted of all individuals with CMT. pLoF variants were exclusively found within Groups 1 and 3, and NLS variants were exclusively seen in Group 2. The distinction between Groups 1 and 3 is not immediately apparent but may reflect phenotypic depth (average number of HPO terms per group: Group 1, 13.00 ± 5.16; Group 2, 22.33 ± 5.51; Group 3, 3.25 ± 2.06; Group 4, 9.67 ± 2.52).

### Functional characterization of DHX9 variant alleles

We next investigated for potential cellular phenotypes and studied whether *DHX9* variants identified in patients affect protein subcellular localization in human cells. WT DHX9 protein exhibits diffuse nuclear expression (Human Protein Atlas, https://www.proteinatlas.org/ENSG00000135829-DHX9). Consistently, we observed EGFP-tagged WT DHX9 protein diffusely distributed in the nucleus of MCF-7 cells (**Fig. 5A**). In contrast, the EGFP-tagged NLS variant p.(Lys1163Arg) resulted in mutant protein localization solely within the cytoplasm (**Fig. 5A**). This disruption of DHX9 nuclear localization was also observed in fibroblasts derived from patient 5 with the p.(Lys1163Arg) variant but not in his unaffected father (**Fig. 5B**). A comparable cytoplasmic distribution was also observed for the NLS variant p.(Arg1166Pro) (**Fig. S5**). In contrast, nonsense or frameshift variants identified in mild NDD patients resulted in EGFP-tagged mutant protein in both the nucleus and cytoplasm (**Fig. 5A**, **Fig. S5**). Missense variants identified in patients with CMT2 instead demonstrated prominent and uniform nucleolar localization as evidenced by their co-staining with the nucleolus marker Fibrillarin (FBL) (**Fig. 5A**, **Fig. S5**). Curiously, cells transfected with mild NDD-associated variants p.(Gly411Glu), p.(Val473Ile), p.(Cys608Gly), and p.(Arg761Gln) showed either diffuse nuclear or nucleolar patterns (**Fig. S6, Table S2**). We further confirmed the localization pattern changes in another human cell line (PC-3) for representative DHX9 mutant proteins (**Fig. 5A**).

**Figure 5:**
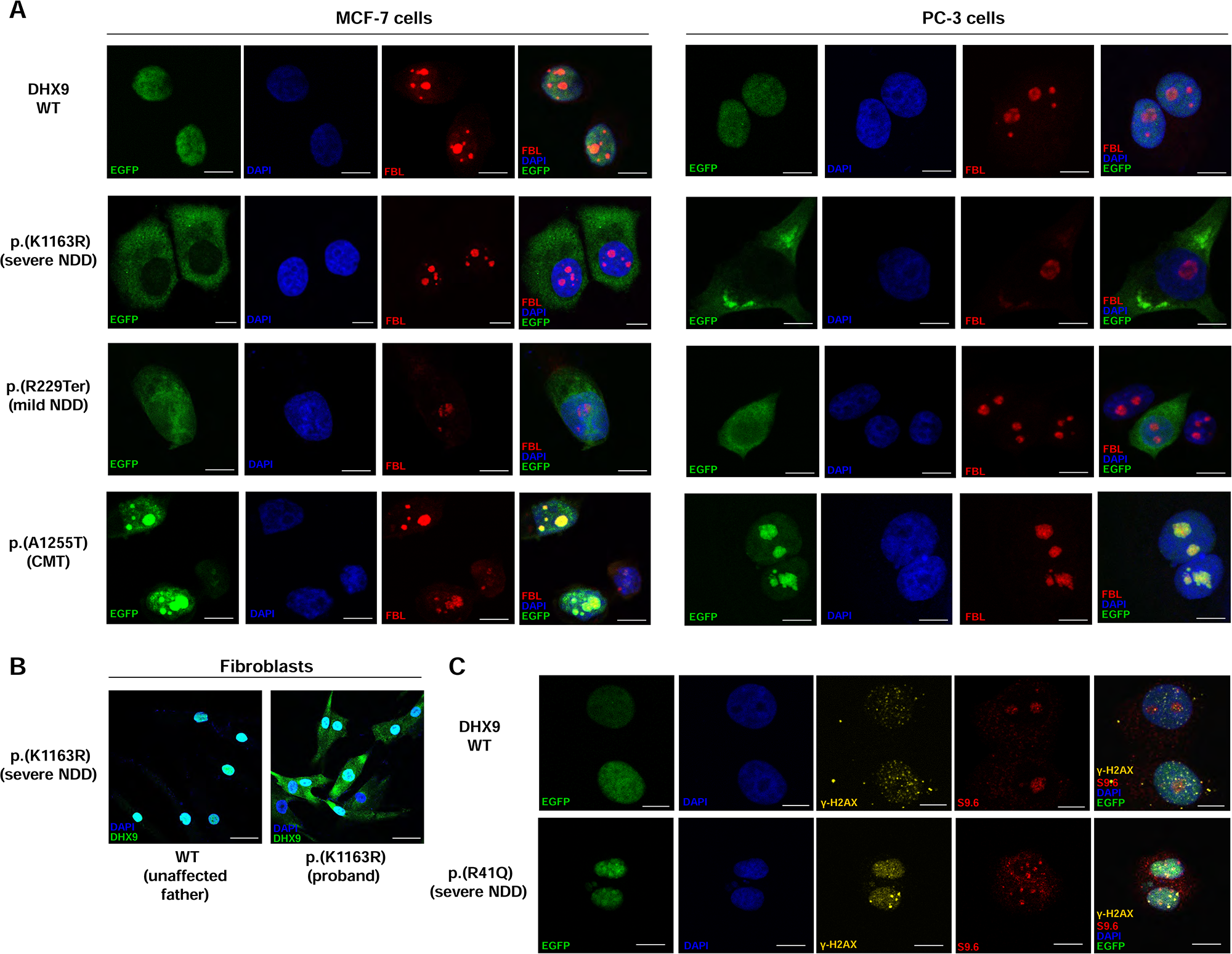
Subcellular localization of DHX9 WT and representative mutant proteins. (A) Subcellular localization of EGFP-tagged DHX9 WT protein, NLS p.K1163R (severe NDD), truncating p.R229* (mild NDD) and CMT p.A1255T mutant proteins expressed in MCF-7 and PC-3 human cells. Nucleolar loci were co-stained by the Fibrillarin (FBL) marker and DNA stained by DAPI. Scale bar = 10 µm. (B) Endogenous expression of DHX9 in fibroblasts from the severe NDD proband with the p.K1163R NLS variant and the unaffected father. Scale bar = 40 µm. (C) Staining of levels of R-loop formation by the S9.6 marker and DSB by the γ-H2AX marker in MCF-7 cells expressing WT or the p.R141Q mutant protein. Scale bar = 10 μm. See also Figure S5, S6.

We also examined the R-loop and DSB levels in cells expressing DHX9 mutant proteins by immunofluorescence (IF) staining of the S9.6 and γ-H2AX markers, respectively. Cells expressing empty vector showed low levels of R-loops and DSBs (**Fig. S5**). The DHX9 p.(Arg141Gln) variant associated with severe NDD induced high levels of R-loops and DSBs with ubiquitous staining throughout the nucleus; in contrast, cells expressing WT DHX9 protein exhibited low levels of DSBs and moderate levels of R-loops (**Fig. 5C**). Truncated DHX9 proteins and NLS mutant proteins induced moderate levels of R-loop and DSB, while cells expressing mild NDD and CMT-associated variants had low levels of R-loops and DSB (**Fig. S5, S6, Table S2**).

Given DHX9 relies on ATP hydrolysis to unwind nucleic acid structures, we inspected whether variants located within helicase domains affect DHX9 ATPase activity. Six variants fall within the helicase ATP-binding or helicase C-terminal domain; these functional domains contain eight conserved motifs (**Fig. S7A**). Among the six variants, the pLoF variants p.(Glu693GlyfsTer7) and p.(Arg764Ter) caused protein truncation and generated baseline ATPase activities comparable to negative controls as expected (**Fig. S7B**). Similar results were obtained for the p.(Arg229Ter) mutant protein with predicted truncation of ATP-binding domains (**Fig. S7B**). Two missense changes located within conserved ATP binding and hydrolysis motifs, p.(Gly411Glu) in Motif I and p.(Arg761Gln) in Motif VI, significantly altered ATPase activity relative to WT DHX9 protein. In contrast, two missense variants located outside of conserved motifs, p.(Val473Ile) and p.(Cys608Gly), demonstrated ATPase activity comparable to WT DHX9 (**Fig. S7B**).

### Dhx9^-/-^ mice are viable but have behavioral and neurological abnormalities

To further explore DHX9’s role in the mammalian nervous system, we generated *Dhx9*^-/-^ mice. Biallelic disruption of *Dhx9* did not impact viability but clearly altered behavioural and neurological function in young adult mice. On exposure to a novel, mildly stressful environment (20-minute open field (**Fig. 6A,B**), introduction to a new home cage (**Fig. 6C**)), *Dhx9*^-/-^ mice exhibited decreased locomotor activity (both forward and vertical) and locomotor speed. Conversely, in a familiar home cage environment, *Dhx9*^-/-^ mice showed increases in vertical locomotor activity during the active dark period of the light/dark cycle (**Fig. 6C**). These abnormal behavioural reactions to different environmental conditions may indicate altered sensory information processing. While both 2-paw and 4-paw grip strength was clearly decreased in *Dhx9*^-/-^ mice (**Fig. 6D**), this was significantly predicted by body weight in linear regression analysis (**Fig. 6E**). In addition, 20% of *Dhx9*^-/-^ mice (3 of 15 mutants, 2 of 6 males, 1 of 9 females) showed tremors in SHIRPA analysis (**Fig. 6F**). The constitutive loss of *Dhx9* also caused deafness in mice as evidenced by absent hearing curves in ABR analysis (**Fig. 6G**) and further reflected by clearly impaired acoustic startle reactivity (**Fig. 6H**). *Dhx9*^-/-^ mice not only have reduced body weight compared with wild-type mice (**Fig. 6I**) but also have reduced food intake and more weight loss than wild-type mice in the 21 h period after transfer to a new home cage (**Fig. 6J, K**).

**Figure 6:**
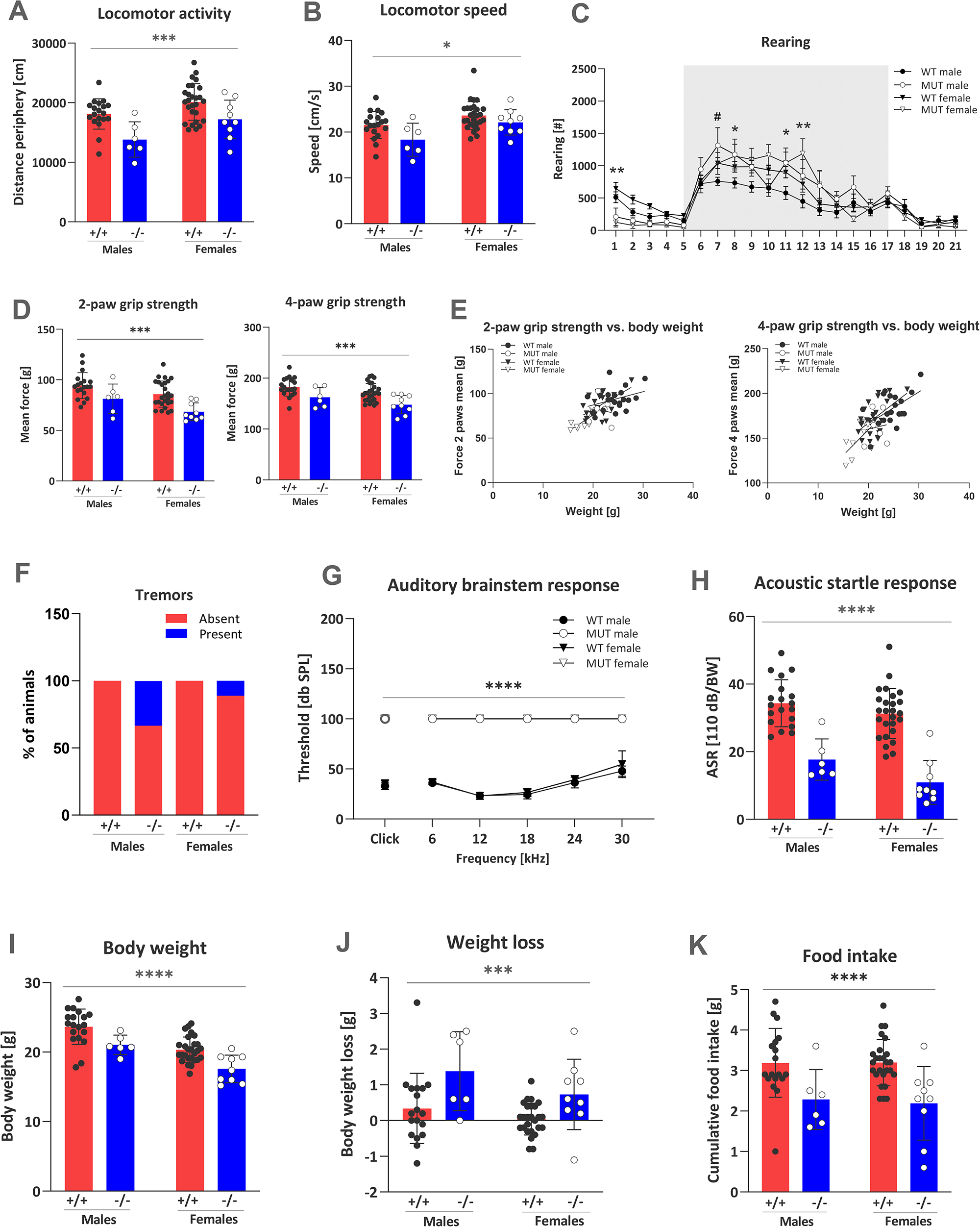
Disruption of *Dhx9* in mice causes behavioral and neurological abnormalities. (A-C) *Dhx9*^-/-^ mice were hypoactive in response to a novel environment (decreased distance travelled (A) and speed (B)) in open field and decreased rearing (C) on first introduction to a novel home cage. However, male *Dhx9*^-/-^ mice showed more rearing activity with lights off during the active phase in the home cage. This increased vertical activity was evident in both male and female *Dhx9*^-/-^ in the latter half of the active phase. Shaded areas encompass period between lights off/lights on, 18:00-06:00, and are mean ± SEM. (D, E) Both 2-paw and 4-paw grip strength were reduced in *Dhx9*^-/-^ mice (D). Lower grip strength was predicted by the lower body weight in linear regression analysis (E). (E) Twenty per cent of *Dhx9*^-/-^ mice showed tremors in the SHIRPA analysis. G, H) Both absent auditory response curve in the auditory brainstem response (G) and decreased acoustic startle reactivity (H) indicated deafness in *Dhx9*^-/-^ mice. (I-K) The body weight of *Dhx9*^-/-^ mice was lower than wild-type mice (I). *Dhx9*^-/-^ mice had lower cumulative food intake and lost more weight over a 21 h period after transfer to a new home cage than wild-type mice. #p < 0.05, male +/+ vs. -/-, *p < 0.05, **p < 0.01, ***p < 0.001, ****p < 0.0001 +/+ vs. -/-. Data are mean ± SD.

In addition to differences in body weight, we observed further indications of altered metabolism and organ function associated with DHX9 loss. Impaired glucose clearance in the ipGTT (**Fig. S8A**) as well as decreased *ad libitum* fed glucose (**Fig. S8B**) and cholesterol levels (**Fig. S8C**) suggest effects on glucose and fat metabolism possibly due to altered liver metabolism. In females, there were increased creatinine and urea levels (**Fig. S8D-E**) suggesting possible altered renal function. Hematologic analysis provided evidence of abnormal erythropoiesis and thrombopoiesis. This included a mild hypochromic microcytosis of erythrocytes as indexed by increased red blood cell counts with decreased mean corpuscular volume (MCV) and mean corpuscular haemoglobin (MCH). Moreover, there were lowered platelet counts in males associated with higher mean platelet volumes and increased anisocytosis of platelets (**Fig. S9**).

## Discussion

Rapid repair of single- and double-strand DNA breaks (SSB and DSB, respectively) is essential for neurodevelopment and neuronal homeostasis^54, 55^. SSBs result from oxidative stress, abortive topoisomerase 1 (Topo1) activity, and as intermediates during base excision repair. They are the most frequently occurring form of DNA damage^55^. Replication-induced DSBs occur during neurogenesis and are repaired by homologous recombination (HR), non-homologous end-joining (NHEJ), and FoSTeS/MMBIR (fork stalling and template switching/microhomology-mediated break-induced replication) at collapsed forks resulting in oeDNA (one ended, double stranded, DNA breaks) due to DNA replication through a nick^15^, whereas transcription-induced DSBs occur in postmitotic neurons and are repaired through NHEJ^54^ and potentially MMBIR.

While DSBs cause genomic instability, compromise genome integrity, and trigger apoptosis, they also play important physiologic roles in the nervous system. For example, neuronal stimulation triggers DSBs in early-response genes, and DSBs trigger early-response gene expression in a stimulus-independent fashion^56^. Similarly, novel environment exploration in mice increased DSBs in the dentate gyrus, a brain region involved in learning and memory^57^. DSBs may contribute to the CNVs detected in healthy human cortices and somatic mosaicism in the developing brain^54, 58^.

Although many key players and mechanisms in SSB/DSB repair have been identified through model organism research and *in vitro* studies, the study of Mendelian DNA repair disorders provides unique insight into SSB/DSB repair’s role in human development, human organismal biology, and disease. For example, spinocerebellar ataxia with axonal neuropathy 1 (SCAN1, [MIM: 607250]) is an adult-onset AR neurological disease trait due to variants in *TDP1*, the gene encoding tyrosyl-DNA phosphodiesterase 1^14(p1)^. Identified in a single consanguineous family, *TDP1* p.(His493Arg) disrupts the phosphodiesterase active site, resulting in catalytically inactive SSB repair complexes and increased SSBs in response to inhibited topoisomerase 1 activity or oxidative stress^14(p1),15, 59(p1),60^. These studies demonstrated for the first time SSB repair’s importance for the maintenance and survival of postmitotic neurons in the cerebellum and peripheral nervous system^12, 13^. Since the recognition of SCAN1 as neurodegenerative disorder resulting from SSB repair over 20 years ago, Mendelian gene discoveries in other neurodegenerative or neurodevelopmental disorders have identified additional genes and pathways involved in SSB/DSB repair, expanding our understanding of their involvement in the human nervous system, health, and disease^55(p),61–65^.

Recent evidence indicates a role for the nuclear protein DHX9 in SSB/DSB prevention and repair. First, DHX9 regulates R-loops, triple-stranded DNA-RNA hybrids which form during transcription^10, 66^. R-loops are abundant in transcriptionally active, GC-rich promoters where they inhibit DNA methylation and therefore stimulate gene expression^11, 13^. However, they are also a source of SSB and DSB: SSB from displaced and exposed single-stranded DNA and DSB from replication fork stalling^11^. While DHX9 was demonstrated to resolve R-loops in some studies^10^, others have shown the opposite^66(p9)^, perhaps indicating context-specific roles.

Second, DHX9 is also essential for DSB repair via HR^12(p9)^. In a recent model, DSB repair is initiated by DHX9’s recruitment to DSBs in transcriptionally active regions in an RNA- and RNA polymerase II-dependent fashion^12(p9)^. Upon migrating to DSBs, DHX9 binds and recruits BRCA1. The DHX9-BRCA1 complex subsequently inhibits 53BP1-mediated NHEJ and recruits HR proteins CTIP, MRE11, and BLM. Consistent with these studies, we observe an increase in R-loops and DSB in human cell lines transfected with DHX9 variant alleles.

These mechanistic studies are complemented by model organism data demonstrating DHX9’s critical involvement in development and biological homeostasis consistent with health. For example, an initial *Dhx9* knockout mouse line suggested DHX9 was indispensable during embryonic development^67^. *Dhx9*^-/-^ mouse embryos were recovered at E10.5 but not E16.5 and exhibited abnormal gastrulation with apoptosis of embryonic ectodermal cells^67^. Similarly, the *Drosophiia* ortholog *mle* (*maleless*) is required for chromosome X dosage compensation^68^ and exhibits distinct LoF and gain-of-function (GoF) phenotypes: homozygous *mle* LoF alleles cause male-specific lethality^68^, whereas the homozygous *mle* GoF allele *nap* causes temperature-sensitive paralysis, decreased lifespan, and neurodegeneration^69, 70^. The *nap* allele alters splicing of *para* (*paralytic*), a voltage-gated sodium channel, and results in decreased sodium channel abundance^71^. Despite these observations, the *DHX9*-associated disease trait(s) in humans have remained elusive.

Here, we provide evidence for the existence of at least two autosomal dominant (AD) *DHX9*-associated rare disease traits: DD/ID and axonal CMT2. Like *DDX3X* and *DHX30*, the best characterized DDX/DHX disease gene paralogs^3–5,6(p30),44(p30)^, *de novo DHX9* missense and pLoF variant alleles associate with a wide phenotypic spectrum. Among patients with NDDs, the spectrum ranged from neurobehavioral differences of isolated ASD without ID on one end to severe DD/ID with microcephaly, brain anomalies, refractory epilepsy, and movement disorders on the other.

Other parallels with *DDX3X* and *DHX30* include the association between structural brain abnormalities with severe phenotypes and variable but frequent extra-neurological findings. Quantitative HPO analysis of the *DHX9* cohort demonstrated four distinct phenotypic clusters: two associated with mild DD/ID (Groups 1 and 3), one associated with severe DD/ID (Group 2), and another associated with CMT (Group 4). While the total number of cases was modest (n=14), these analyses provide evidence consistent with the contention of emerging genotype-phenotype correlations reminiscent of DDX3X and DHX30: an association of pLoF variant alleles with mild NDD phenotypes and specific missense variants with severe phenotypes^3–5,6(p30),44(p30)^. The sole case of a maternally inherited 191.52 kb CNV deletion encompassing *DHX9* in DECIPHER in an individual with a neurodevelopmental phenotype may corroborate this genotype-phenotype schema; alternatively, it may indicate reduced penetrance of *DHX9* LoF. Additional case ascertainment and variant studies are needed to further explore these hypotheses.

The association of a single gene with distinct phenotypes such as NDDs and CMT2 may be surprising but is not without precedence. The allelic series of *PNKP*, a gene encoding another SSB/DSB repair enzyme, polynucleotide kinase 3-prime phosphatase, is relevant as it ranges from Charcot-Marie-Tooth disease, type 2B2 [MIM: 605589] to ataxia-oculomotor apraxia 4 [MIM: 616267] and microcephaly, seizures, and developmental delay [MIM: 613402]^72^. Other examples include *SURF1* (CMT type 4K [MIM: 616684]; mitochondrial complex IV deficiency, MIM: 220110), *YARS1* (CMT, dominant intermediate C [MIM: 608323]; infantile-onset multisystem neurologic, endocrine, and pancreatic disease 2 [MIM: 619418]), *ATP1A1* (CMT, axonal, type 2DD [MIM: 618036]; hypomagnesemia, seizures, and mental retardation 2 [MIM: 618314]), *MORC2* (CMT, axonal, type 2Z [MIM: 616688]; developmental delay, impaired growth, dysmorphic facies, and axonal neuropathy [MIM: 619090]), and *AIFM1* (combined oxidative phosphorylation deficiency 6 [MIM: 300816]; Cowchock syndrome [MIM: 310490]). As *PNKP*, *SURF1*, *YARS1*, *ATP1A1*, *MORC2*, *AIFM1*, and *DHX9* all play integral roles in neuronal homeostasis and survival, each gene’s phenotypic spectrum likely reflects distinct mutational mechanisms within the allelic series (*e.g.*, amorphs versus hypomorphs) and a phenotypic gradient caused by a range of residual gene function or gene dosage^73^.

The mechanisms underlying *DHX9*-related neurologic disorders are likely complex and will take a great deal of experimental biology, human genomics, and time to dissect. The paucity of pLoF variants in gnomAD and the exclusive identification of high confidence pLoF variants in diseased individuals in research and clinical genomic databases suggests haploinsufficiency as a disease mechanism as in *DDX3X* and *DHX30*. However, the increased R-loop and DSBs induced by *DHX9* variants p.(Arg141Gln), p.(Arg1166Pro), and p.(Lys1163Arg) and their association with severe phenotypes points towards a gain-of-function (GoF), perhaps dominant negative mechanism. Similarly, the nucleolar accumulation of DHX9 seen in CMT- and mild NDD-associated *DHX9* variant might indicate a separate, distinct GoF mechanism. As cellular stressors including RNA PolII-mediated transcriptional inhibition, growth arrest, viral replication, or hypothermia all induce nucleolar DHX9 translocation^74, 75^, it is possible that pathogenic *DHX9* variants may disrupt neurodevelopment or cause neurodegeneration by inducing cellular stress responses.

Consistent with pathogenic *DHX9* variant alleles association with DD/ID and CMT in humans, we find homozygous deletion of the mouse ortholog *Dhx9* results in multiple behavioral and neurological abnormalities (**Fig. 6**). Although further phenotyping of aged mice is needed, the observation of tremor in a subset of young adult mice is intriguing as Trembler and *Trembler^J^* mice, both *Pmp22* CMT models, also exhibit tremor^76, 77^. Curiously, the homozygous null line described here was viable in contrast to the previously reported knockout mouse line which exhibited embryonic lethality^67^. While the explanation for the variation in viability is not currently clear, genetic construct and background differences may play a role. An important construct distinction in the Lee *et al*. 1998 model was the insertion of a neomycin resistance (neo) cassette in the opposite reading frame in the middle of exon 2. On the C57BL/6 (unspecified substrain) x Sv129 mixed background, the neo cassette, may have been toxic. This also illustrates how genetic modifiers and epigenetic variation between different genetic backgrounds may modify how mutagenesis approaches influence the essentialome^78, 79^.

In summary, we provide evidence of *DHX9* as a cause of two AD human neurogenetic disease traits: DD/ID and axonal CMT. Our allelic affinity studies, like those performed on *SOX10* with nonsense variants/premature termination codons (PTCs) conveying different neurological disease traits depending upon whether they escape nonsense mediated decay (PCWH syndrome; [MIM: 609136]) or not (Waardenburg syndrome, type 4C; [MIM: 613266]), provide insight into human nervous system development^73, 80^. Our allelic series provides a starting point for the dissection of the role of *DHX9* in human neurodevelopment and neurodegeneration. As in *DDX3X* and *DHX30*, expansion of the *DHX9* allelic series and functional studies will facilitate genotype-phenotype correlations and provide greater insights into the contribution of *DHX9* to human health and disease.

## Data Availability

All data produced in the present study are available upon reasonable request to the authors

---

**Supplemental Figure 1. Phenotyping pipeline used to analyse *Dhx9* -/- mice showing categories and tests performed and mouse age in weeks at testing**

**Supplemental Figure 2: DHX9 protein – protein interactome**

Interactome data taken from STRING database (https://string-db.org/).

**Supplemental Figure 3: Analysis of nuclear localization signal variants**

(A) Conservation of all amino acids which fall within the NLS. Lys1163 and Arg1166 are highlighted.

(B) cNLS Mapper prediction for reference sequence. Red letters indicate predicted NLS. Yellow highlight indicates known NLS.

(C) cNLS prediction for p.(Lys1163Arg) variant sequence. A NLS was not identified.

(D) cNLS prediction for p.(Arg1166Pro) variant sequence. A NLS was not identified.

**Supplemental Figure 4: *In silico* analysis of *DHX9* variant c.627-4dupA**

(A) Diagram demonstrating impact of *DHX9*: c.627-4dupA on splicing junction.

(B) *In silico* prediction from Human Splice Finder (http://www.umd.be/hsf/)

(C) *In silico* prediction from Splice AI (https://spliceailookup.broadinstitute.org/).

**Supplemental Figure 5. Subcellular localization of DHX9 mutant proteins and the levels of R-loop and DNA damage.** Scale bar = 10 µm. See also Figure 5

**Supplemental Figure 6. Subcellular localization of remaining DHX9 mutant proteins and the levels of R-loop and DNA damage.** Scale bar = 10 µm.

**Supplemental Figure 7. DHX9 missense variants located within the ATP binding and hydrolysis conserved motifs affected ATPase activity.**

(A) Schematic representation of patients’ variants with regards to functional domains of the DHX9 protein. Amino acid sequence of the helicase ATP-binding and helicase C-terminal domains of DHX9 is listed. The eight conserved motifs of these functional domains are marked within boxes. Corresponding sequence of DHX8 is aligned together to demonstrate the high conservation of the eight motifs. Truncating variants (R229*, E693Gfs*7 and R764*) are labeled in orange, missense variants located within conserved motifs (G411E and R761Q) are labeled in dark blue, and the remaining missense variants (V473I and C608G) are labeled in light blue.

(B) Assays demonstrated the relative ATPase activities of DHX9 mutant proteins with various missense changes compared to WT protein. In each experiment, ATPase activity was normalized to the amount of purified protein. A representative Coomassie blue staining image demonstrating the sizes and expression levels of purified DHX9 proteins is shown here. Experiments were repeated at least three times for each DHX9 variant. See Table S3 for raw data on absorbance values of each sample. Note that for truncating variants (R229*, E693Gfs*7 and R764*), the absorbance values were comparable to the baseline (no transfection blank and EGFP backbone only expression), therefore, their relative ATPase activities to WT protein were not calculated. Also note that for the p.R761Q protein, its much lower expression level relative to WT caused its higher calculated ATPase activity, given the calculation was normalized based on corresponding protein amount. Its actual ATPase activity values (raw data on absorbance values) were consistently lower than the WT values (Table S3). **, p<0.005; *, p<0.05; One-Way ANOVA.

**Supplemental Figure 8. Loss of *Dhx9* in mice causes differences in clinical chemistry indices indicative of altered metabolism and renal function.**

**Supplemental Figure 9. Loss of *Dhx9* in mice causes haematological alterations indicative of effects on erythropoiesis and thrombopoiesis**.

**Table S1. Number of *Dhx9 -/-* tested in the assays where relevant differences were detected**.

**Table S2. Summary on disease type and severity, subcellular localization, R-loop and DNA damage levels in cells expressing various DHX9 proteins.**

WT, wildtype; NLS, nuclear localization signal domain; NDD, neurodevelopmental diseases; CMT, Charcot-Marie-Tooth disease; DSB, double-stranded break.

**Table S3. Raw data on absorbance values of ATPase activity experiments**.

## Web Resources

Online Mendelian Inheritance in Man, http://www.omim.org

gnomAD Browser, https://gnomad.broadinstitute.org/

Baylor College of Medicine Human Genome Sequencing Center, https://www.hgsc.bcm.edu

Baylor College of Medicine Lupski Lab, https://github.com/BCM-Lupskilab

Genotype-Tissue Expression (GTEx), https://gtexportal.org/home/

BrainSpain, https://www.brainspan.org/

UCSC Cell Browser, https://cells.ucsc.edu/

STRING, https://string-db.org/

Human Splice Finder, https://www.genomnis.com/access-hsf

CADD, https://cadd.gs.washington.edu/

## Data Availability

All data described in this study are provided within the article and Supplementary Material. Raw sequencing data and de-identified clinical data are available from the corresponding authors upon request.

## Acknowledgements

This study was supported in part by the U.S. National Human Genome Research Institute (NHGRI) and National Heart Lung and Blood Institute (NHBLI) to the Baylor-Hopkins Center for Mendelian Genomics (BHCMG, UM1 HG006542, J.R.L); NHGRI grant as part of the GREGoR Consortium (U01 HG011758 to J.E.P., J.R.L., and R.A.G.); NHGRI grant to Baylor College of Medicine Human Genome Sequencing Center (U54HG003273 to R.A.G.); U.S. National Institute of Neurological Disorders and Stroke (NINDS) (R35NS105078 to J.R.L), Muscular Dystrophy Association (MDA) (512848 to J.R.L.), and Spastic Paraplegia Foundation Research Grant to J.R.L. This study was also supported by the General Research Fund from Research Grants Council of Hong Kong (24101921 to S.G.) and National Natural Science Foundation of China (82202045 to S.G.). D.M. was supported by a Medical Genetics Research Fellowship Program through the United States National Institute of Health (T32 GM007526-42). T.M. is supported by the Uehara Memorial Foundation. D.P. was supported by a NINDS 1K23 NS125126-01A1 and Rett Syndrome Research Trust fellowship award from International Rett Syndrome Foundation (IRSF grant #3701-1). J.E.P. was supported by NHGRI K08 HG008986. D.G.C. was supported by NIH – Brain Disorders and Development Training Grant (T32 NS043124), NIH Medical Genetics Research Fellowship Program (T32 GM007526), the Chao Physician Scientist Award, and MDA Development Grant (873841). This study was supported by the CHU de Dijon Bourgogne and by a grant from the French Ministry of Health «DIS-SEQ – Evaluation médico-économique des différentes stratégies de technologies de séquençage par haut débit dans le diagnostic des patients atteints de déficience intellectuelle», Clinical Trial NCT03287206. The Deciphering Developmental Disabilities (DDD) study presents independent research commissioned by the Health Innovation Challenge Fund [grant number HICF-1009-003], a parallel funding partnership between Wellcome and the Department of Health, and the Wellcome Sanger Institute [grant number WT098051]. The views expressed in this publication are those of the author(s) and not necessarily those of Wellcome or the Department of Health. The study has UK Research Ethics Committee approval (10/H0305/83, granted by the Cambridge South REC, and GEN/284/12 granted by the Republic of Ireland REC). The research team acknowledges the support of the National Institute for Health Research, through the Comprehensive Clinical Research Network. The work by Tomi Pastinen and Isabelle Thiffault was made possible by the generous gifts to Children’s Mercy Research Institute and Genomic Answers for Kids program at Children’s Mercy Kansas City. Tomi Pastinen holds the Dee Lyons/Missouri Endowed Chair in Pediatric Genomic Medicine. Research reported in this manuscript was supported by the NIH Common Fund, through the Office of Strategic Coordination/Office of the NIH Director under Award Number(s) [UO1HG007672-Shashi]. The content is solely the responsibility of the authors and does not necessarily represent the official views of the National Institutes of Health.

## Ethics Declaration

This study adheres to the principles in the Declaration of Helsinki. The study was approved by Baylor College of Medicine Institutional Review Board (IRB) protocol H-29697. The work by Tomi Pastinen and Isabelle Thiffault were approved by the Children’s Mercy Institutional Review Board (study # 11120514). Written informed consent was obtained from all participants as required by the IRB. Consent forms are archived and available upon request.

## Potential Conflict of Interest

J.R.L. has stock ownership in 23 andMe, is a paid consultant for Genome International, and is a co-inventor on multiple United States and European patents related to molecular diagnostics for inherited neuropathies, eye diseases, genomic disorders, and bacterial genomic fingerprinting. The Department of Molecular and Human Genetics at Baylor College of Medicine receives revenue from clinical genetic testing conducted at Baylor Genetics (BG) Laboratories. Francisca Millan and Teresa Santiago-Sims are employees of GeneDx. Other authors have no potential conflicts to disclose.

## Notes

### Competing Interest Statement

The authors have declared no competing interest.

### Author Declarations

The study was approved by Baylor College of Medicine Institutional Review Board (IRB) protocol H-29697. The work by Tomi Pastinen and Isabelle Thiffault were approved by the Children's Mercy Institutional Review Board (study # 11120514).

