## Supplemental Figures and Text for "Monoallelic variation in *DHX9*, the gene encoding the DExH-box helicase DHX9, underlies neurodevelopment disorders and Charcot-Marie-Tooth disease"

### Supplemental Figure 1

| | | Age (weeks) | | | | | | | | | |
|---|---|---|---|---|---|---|---|---|---|---|---|
| **Lab/Screen** | Methods | 7 | 8 | 9 | 10 | 11 | 12 | 13 | 14 | 15 | 16 |
| Behaviour | Openfield | | ■ | | | | | | | | |
| | Acoustic startle response & PPI | | | | ■ | | | | | | |
| Neurology | Modified SHIRPA, grip strength | | | ■ | | | | | | | |
| | Rotarod | | | | ■ | | | | | | |
| Dysmorphology | Anatomical observation | | | ■ | | | | | | | |
| Energy Metabolism | Indirect calorimetry | | | | | ■ | | | | | |
| Cardiovascular | Awake ECG / Echo cardiography | | | | | | ■ | | | | |
| Clinical Chemistry | IpGTT | | | | | | | ■ | | | |
| Neurology | Auditory brain stem response (ABR) | | | | | | | | ■ | | |
| Dysmorphology | X-Ray, DEXA | | | | | | | | ■ | | |
| Eye | Scheimpflug imaging, Laser-interference-biometry (LIB), Optical coherence tomography (OCT), Virtual drum test | | | | | | | | | ■ | |
| Clinical Chemistry | Clinical Chemical analysis, hematology | | | | | | | | | | ■ |
| Immunology | Flow cytometry, plasma (IgE, IL6, TNF, insulin) | | | | | | | | | | ■ |
| Pathology | Macro & microscopic analysis | | | | | | | | | | ■ |

**Fig. S1.** Phenotyping pipeline used to analyse *Dhx9* -/- mice showing category and test performed and mouse age in weeks at testing

### Supplemental Figure 2

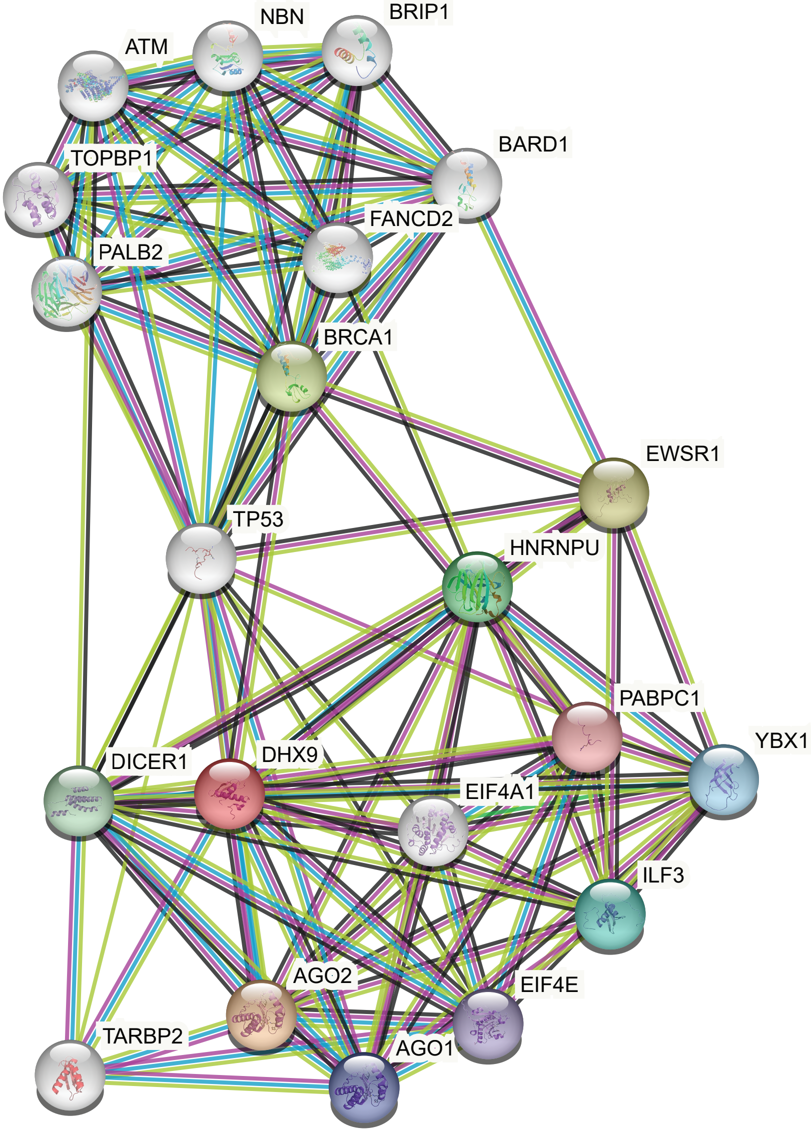

**DHX9 interactome - https://string-db.org/**

### Supplemental Figure 3

A)

| | Nuclear localization signal | | | | | | | | | | | | | | | | | |
|---|---|---|---|---|---|---|---|---|---|---|---|---|---|---|---|---|---|---|
| *H. sapiens* | Y | G | D | G | P | R | P | P | K | M | A | R | Y | D | N | G | S | G | Y |
| *P. troglodytes* | Y | G | D | G | P | R | P | P | K | M | A | R | Y | D | N | G | S | G | Y |
| *M. mulatta* | Y | G | D | G | P | R | P | P | K | M | A | R | Y | D | N | G | S | G | Y |
| *P. hamadryas* | Y | G | D | G | P | R | P | P | K | M | A | R | Y | D | N | G | S | G | Y |
| *B. taurus* | Y | G | D | G | P | R | P | P | K | M | A | R | Y | D | N | G | S | G | Y |
| *E. caballus* | Y | G | D | G | P | R | P | P | K | M | A | R | Y | D | N | G | S | G | Y |
| *C. lupus familiaris* | Y | G | D | G | P | R | P | P | K | M | A | R | Y | D | N | G | G | G | Y |
| *M. musculus* | Y | G | D | G | P | R | P | P | K | M | A | R | Y | D | N | G | S | G | Y |
| *R. norvegicus* | Y | G | D | G | P | R | P | P | K | M | A | R | Y | D | N | G | S | G | Y |
| *L. africana* | Y | G | D | G | P | R | P | P | K | M | A | R | Y | D | N | G | S | G | Y |
| *D. novemcinctus* | Y | G | D | G | P | R | P | P | K | M | A | R | Y | D | N | G | S | G | Y |
| *A. carolinensis* | Y | G | D | G | P | R | P | P | K | M | A | R | Y | D | N | G | G | G | Y |
| *D. rerio* | F | G | D | G | P | R | P | P | K | M | A | R | T | D | F | G | G | G | F |

#### B) WT DHX9

**Predicted NLSs in query sequence**

```
IVLVDDWIKLQISHEAAACITGLRAAMEALVVEVTKQPAIISQLDPVNER  50
MLNMIRQISRPSAAGINLMIGSTRYGDGPRPPKMARYDNGSGYRRGGSSY 100
SGGGYGGGYSSGGYGSGGYGGSANSFRAGYGAGVGGGYRGVSRGGFRGNS 150
GGDYRGPSGGYRGSGGFQRGGGRGAYGTGYFGQGRGGGGY           190
```

| Predicted monopartite NLS | | |
|---|---|---|
| **Pos.** | **Sequence** | **Score** |
| 78 | GPRPPKMARYD | 7.5 |

#### C) p.(Lys1163Arg)

**Predicted NLSs in query sequence**

```
IVLVDDWIKLQISHEAAACITGLRAAMEALVVEVTKQPAIISQLDPVNER  50
MLNMIRQISRPSAAGINLMIGSTRYGDGPRPPRMARYDNGSGYRRGGSSY 100
SGGGYGGGYSSGGYGSGGYGGSANSFRAGYGAGVGGGYRGVSRGGFRGNS 150
GGDYRGPSGGYRGSGGFQRGGGRGAYGTGYFGQGRGGGGY           190
```

| Predicted monopartite NLS | | |
|---|---|---|
| **Pos.** | **Sequence** | **Score** |

#### D) p.(Arg1166Pro)

**Predicted NLSs in query sequence**

```
IVLVDDWIKLQISHEAAACITGLRAAMEALVVEVTKQPAIISQLDPVNER  50
MLNMIRQISRPSAAGINLMIGSTRYGDGPRPPKMAPYDNGSGYRRGGSSY 100
SGGGYGGGYSSGGYGSGGYGGSANSFRAGYGAGVGGGYRGVSRGGFRGNS 150
GGDYRGPSGGYRGSGGFQRGGGRGAYGTGYFGQGRGGGG            189
```

| Predicted monopartite NLS | | |
|---|---|---|
| **Pos.** | **Sequence** | **Score** |

### Supplemental Figure 4

A)

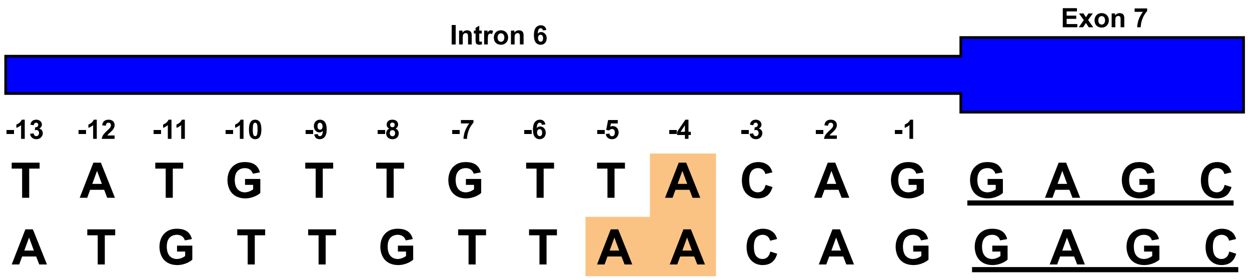

B)

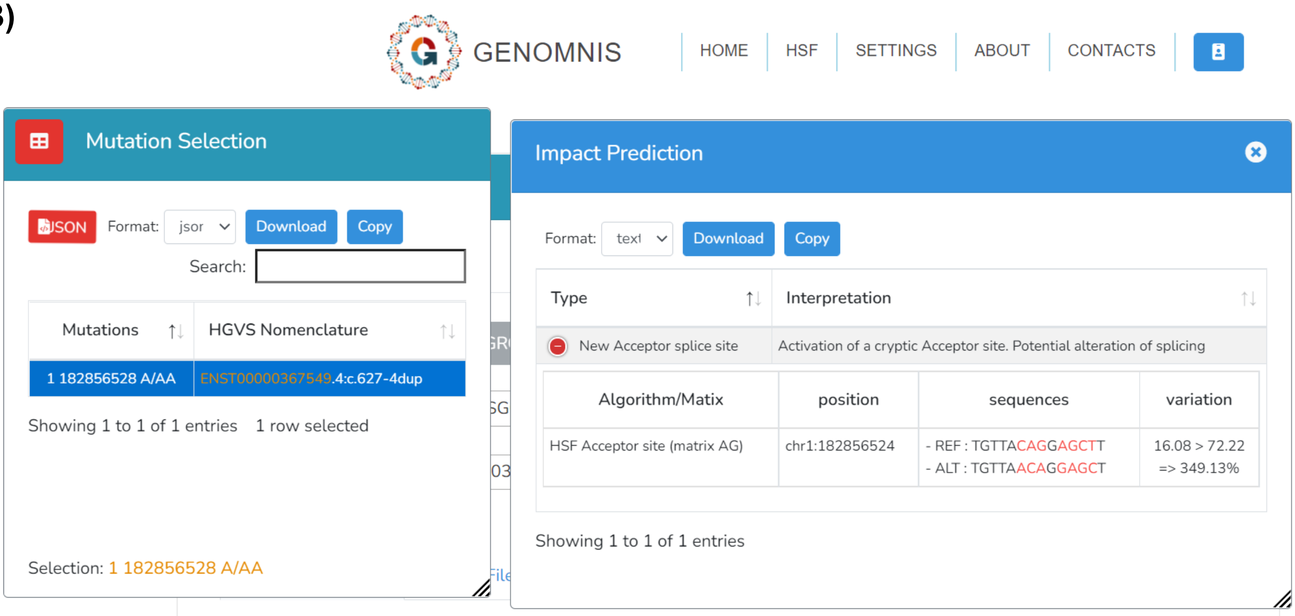

C)

| Δ type | Δ score ⓘ | pre-mRNA position ⓘ |
|---|---|---|
| Acceptor Loss | 0.02 | 4 bp |
| Donor Loss | 0.01 | 50 bp |
| Acceptor Gain | 0.14 | 18 bp |
| Donor Gain | 0.00 | |

### Supplemental Figure 5

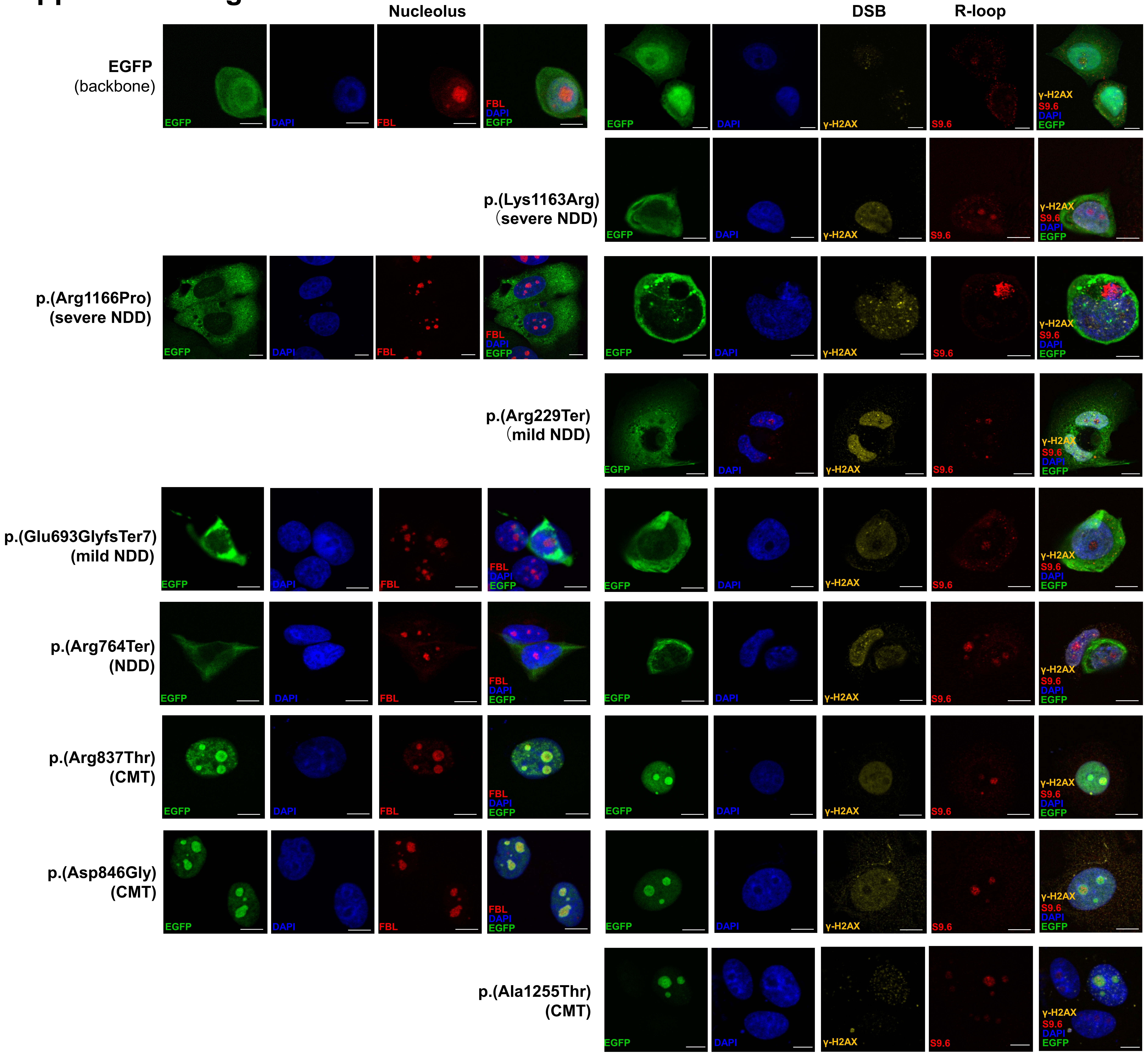

### Supplemental Figure 6

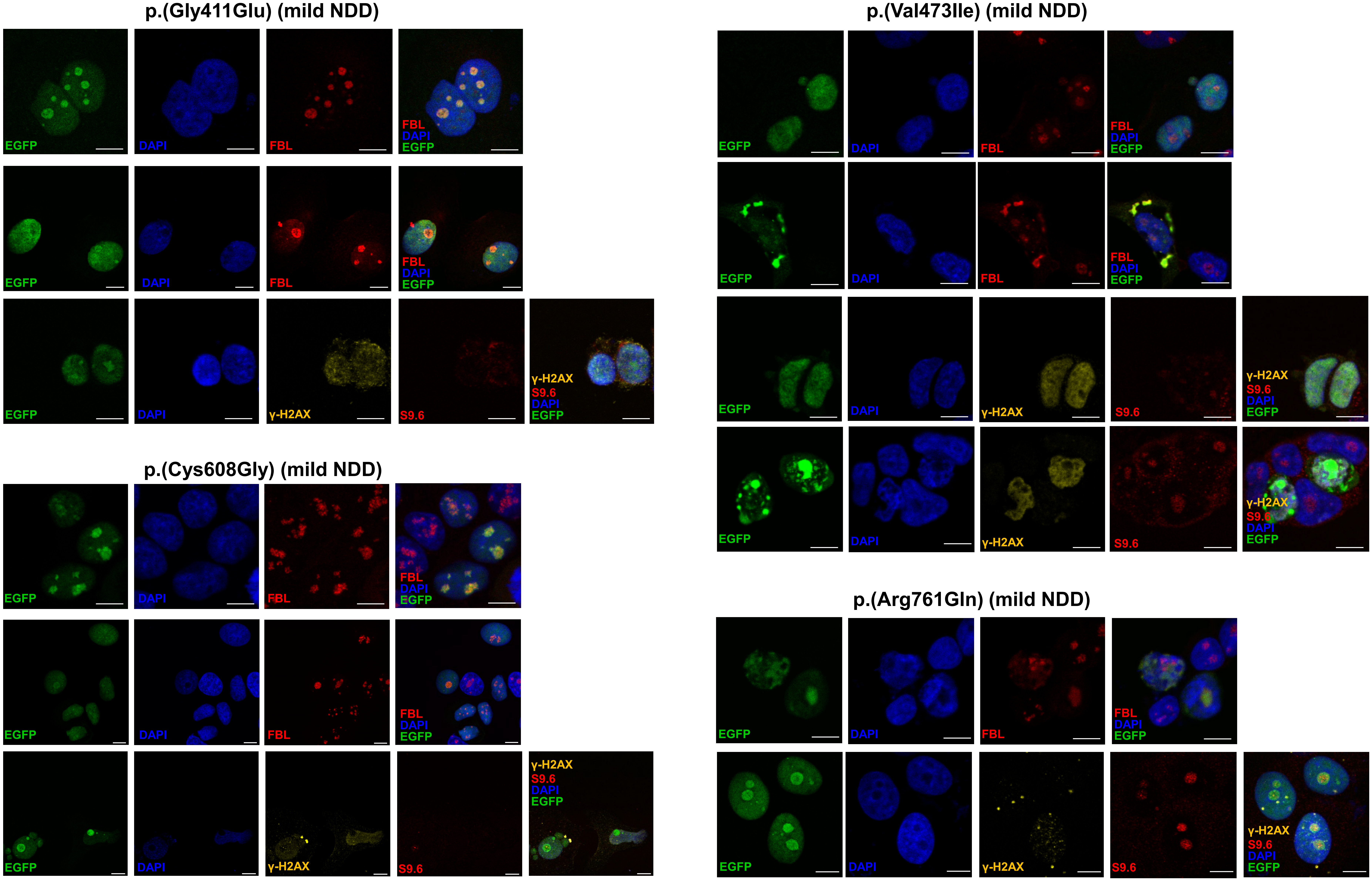

### Supplemental Figure 7

A

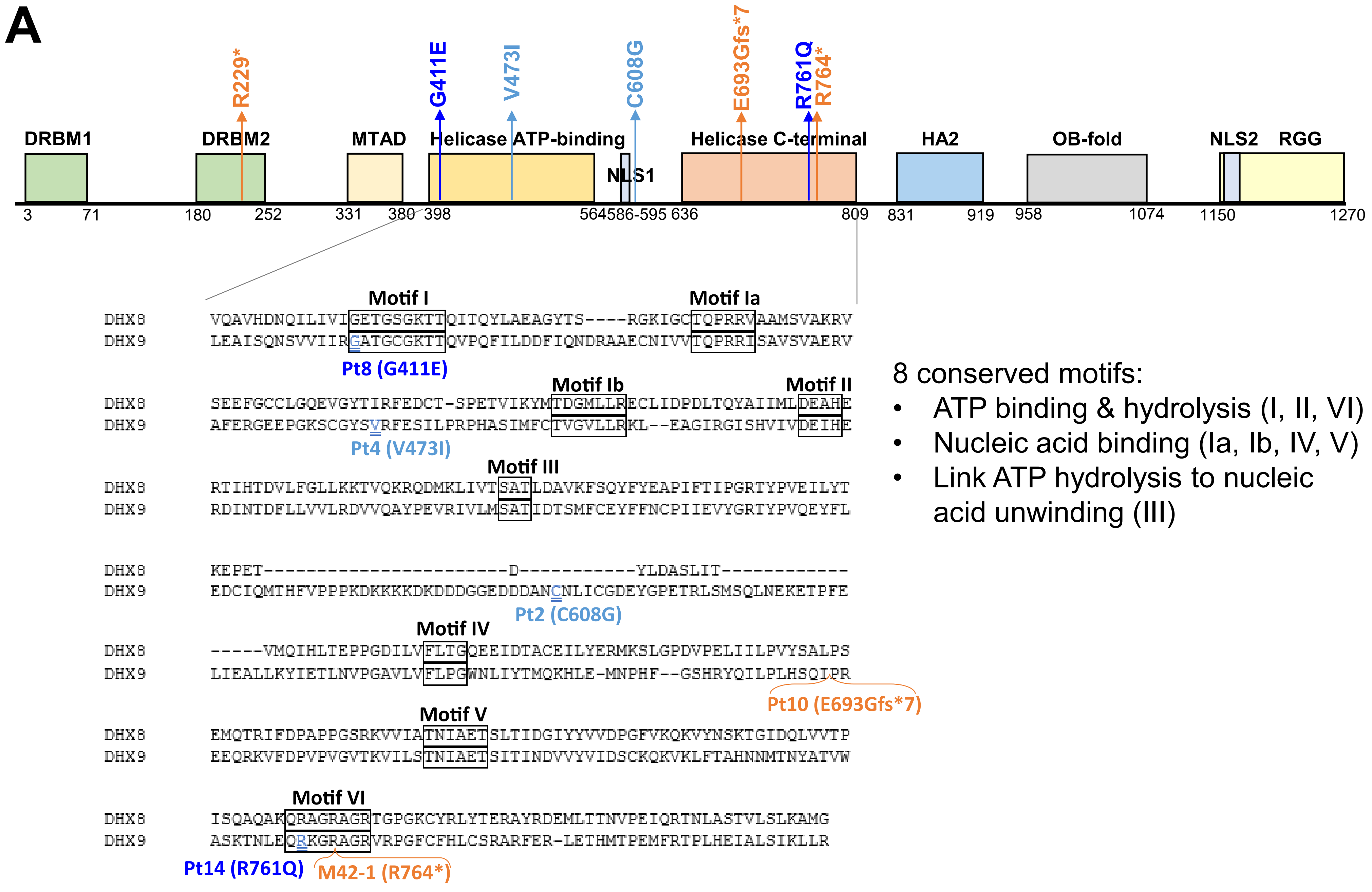

B

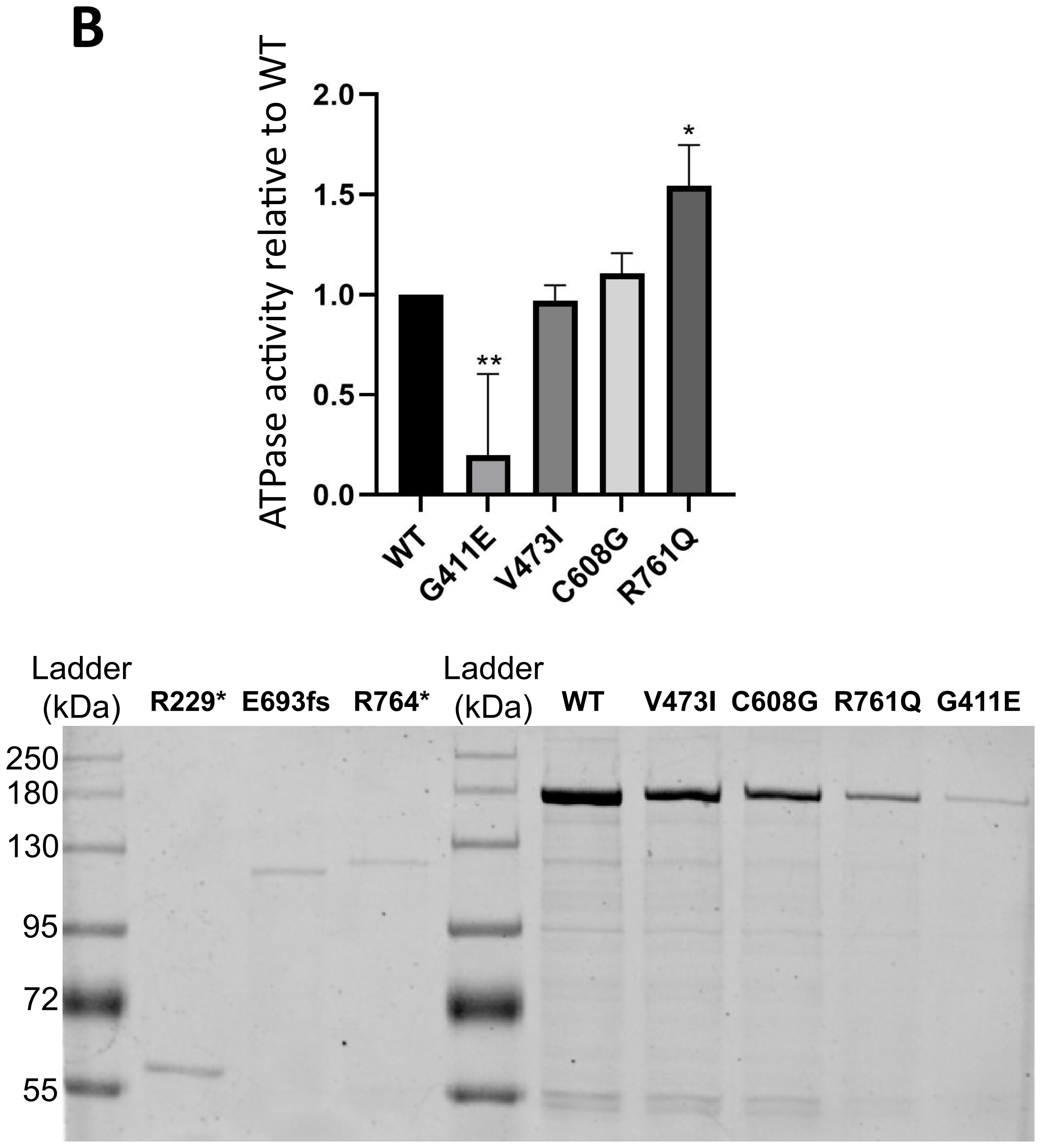

### Supplemental Figure 8

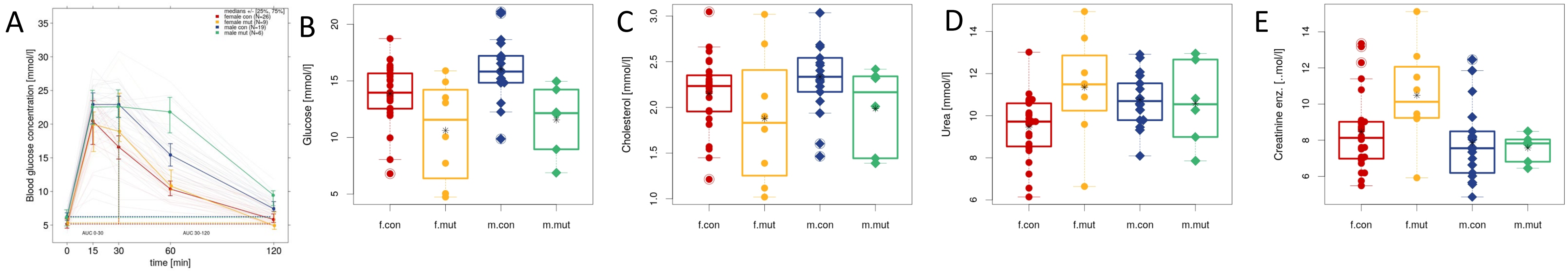

| IpGTT results | female | | male | | linear model | linear model | linear model |
|---|---|---|---|---|---|---|---|
| | con | mut | con | mut | genotype | sex | genotype:sex |
| | n=26 | n=9 | n=19 | n=6 | | | |
| | mean ± sd | mean ± sd | mean ± sd | mean ± sd | p-value | p-value | p-value |
| **Glucose (T=0)** | 5.35 ± 0.81 | 5.42 ± 0.56[a] | 6.23 ± 1.04 | 6.03 ± 1.27 | 0.822 | 0.014 | 0.664 |
| **AUC 0-30** | 295.39 ± 94.35 | 330.05 ± 74.5[a] | 366.05 ± 51.65 | 384.82 ± 65.12[a] | 0.302 | 0.018 | 0.758 |
| **AUC 30-120** | 417.75 ± 176.08 | 551.57 ± 208.3[a] | 724.27 ± 183.29 | 1050.96 ± 226.25[a] | < 0.001 | < 0.001 | 0.123 |

**Fig. S8.** Loss of *Dhx9* in mice causes differences in clinical chemistry indices indicative of altered metabolism and renal function

### Supplemental Figure 9

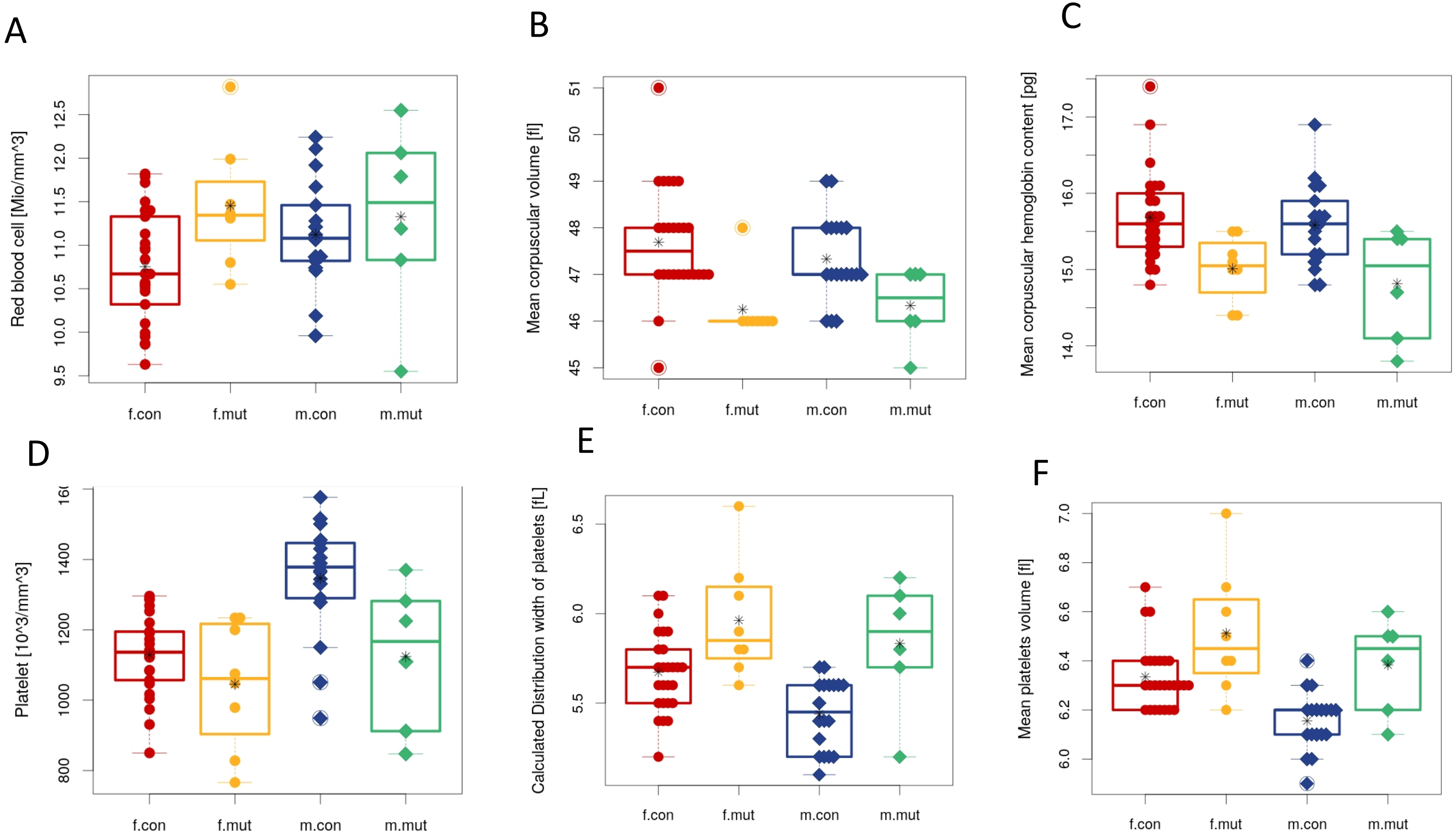

**Fig. S9.** Loss of *Dhx9* in mice causes haematological alterations indicative of effects on erythropoiesis and thrombopoiesis

### Supplemental Table 1

| Assay | Age (weeks) | Number (n) | | | |
|---|---|---|---|---|---|
| | | +/+ | | -/- | |
| | | Males | Females | Males | Females |
| Open field | 8 | 19 | 26 | 6 | 9 |
| SHIRPA | 9 | 19 | 26 | 6 | 9 |
| Grip strength | 9 | 19 | 26 | 6 | 9 |
| Acoustic startle | 10 | 19 | 26 | 6 | 9 |
| Indirect calorimetry | 11 | 18 | 26 | 6 | 9 |
| Glucose tolerance test | 13 | 19 | 26 | 6 | 9 |
| Auditory brainstem response | 14 | 14 | 16 | 4 | 4 |
| Clinical chemistry/hematology | 16 | 19 | 26 | 6 | 9 |

**Table S1.** Number of *Dhx9* -/- tested in the assays where relevant differences were detected.

#### Supplemental Table 2

| Variant Type (Patient ID) | Disease Type | Severity | DHX9 location | DSB pattern and level | R-loop pattern and level |
|---|---|---|---|---|---|
| WT | - | - | Diffuse nuclear | Scattered, low | Gathered, + |
| Truncating (9,10,M42-1) | NDD | Mild | Whole cell | Ubiquitous, + | Gathered, + |
| NLS missense (3,5) | NDD | Severe | Cytoplasm only | Ubiquitous, + | Gathered, + |
| Missense (1) | NDD | Severe | Diffuse nuclear | Ubiquitous, ++ | Ubiquitous, ++ |
| Missense (2,4,8,11) | NDD | Mild | Diffuse nuclear or nucleolar | Ubiquitous, low | Gathered, low |
| Missense (15,16,17) | CMT | - | Nucleolar | Ubiquitous, low | Gathered, low |

### Supplemental Table 3

| | Absorbance at 620nm | | | |
|---|---|---|---|---|
| **Transfected expression plasmid** | Trial 1 | Trial 2 | Trail 3 | Average |
| No transfection blank | 0.442 | 0.441 | 0.441 | 0.441 |
| EGFP-backbone | 0.435 | 0.342 | 0.343 | 0.373 |
| EGFP-DHX9 WT | 0.646 | 0.647 | 0.66 | 0.651 |
| DHX9 p.R229* | 0.377 | 0.433 | 0.417 | 0.409 |
| DHX9 p.G411E | 0.457 | 0.419 | 0.466 | 0.447 |
| DHX9 p.V473I | 0.628 | 0.653 | 0.634 | 0.638 |
| DHX9 p.C608G | 0.648 | 0.688 | 0.656 | 0.664 |
| DHX9 p.E693Gfs*7 | 0.438 | 0.358 | 0.421 | 0.406 |
| DHX9 p.R764* | 0.41 | 0.447 | 0.428 | 0.428 |
| DHX9 p.R761Q | 0.6 | 0.614 | 0.593 | 0.602 |

**Supplemental Text**

Limited clinical details were available for three individuals: two within the BHCMG/GREGoR, BAB4646 and M42-1, and a patient from a simplex autism spectrum disorder (ASD) cohort[19]. BAB4646 and M42-1 have the only two *DHX9* pLoF variants within the BHCMG database of 12,266 exomes and genomes. BAB4646's phenotype is severe DD/ID and primary immunodeficiency. Proband ES identified two heterozygous pathogenic variants in *TRNT1*(NM_182916.2): c.1246A>G, p.(Lys416Glu) and c.608+1G>T. As *TRNT1* causes autosomal recessive syndrome sideroblastic anemia with B-cell immunodeficiency, periodic fevers, and developmental delay [MIM: 616084], these variants likely contribute to the individual's DD/ID and immunodeficiency. Further confirmation of this contention was obfuscated by the lack of additional DNA samples from the proband or his parents precluding variant phasing and determination of *de novo* status.

Subject M42-1 was enrolled in a mitochondrial disease cohort and is one of two affected siblings with encephalopathy, stroke-like episodes, and drug-resistant epilepsy. Proband ES failed to identify a candidate variant to explain the patient's mitochondrial disease, and sibling DNA is not available for testing. While *DHX9* pLoF variants are unlikely to completely explain the phenotypes of BAB4646 and M42-1, they may contribute to their neurologic dysfunction via multi-locus pathogenic variation to a blended traits phenotype[53].
